# Cardiovascular and Thromboembolic Safety Signals Associated with Non-Steroidal Anti-Inflammatory Drugs Initiation: A Sequence Symmetry Analysis

**DOI:** 10.1101/2025.11.26.25341008

**Authors:** Sai Sumedha Bobba, Elin Rowlands, Antonella Delmestri, Wai Yi Man, Xihang Chen, Xintong Li, Anna Saura-Lazaro, Daniel Prieto Alhambra, Danielle Newby

## Abstract

**Background and aims:** Non-steroidal anti-inflammatory drugs (NSAIDs) are widely used, yet real-world evidence on cardiovascular (CV) and venous thromboembolic (VTE) risks for individual agents is limited. We aimed to identify CV and VTE safety signals associated with NSAID initiation.

**Methods:** We conducted a sequence symmetry analysis including patients aged 18+ with one year of prior observation initiating NSAIDs between 2013 to 2023 using CPRD GOLD. Patients had a CV [myocardial infarction (MI), arrythmia, heart failure, haemorrhagic/ischemic stroke] or VTE [deep vein thrombosis (DVT), pulmonary embolism (PE)] within ±180 days of NSAID initiation. Adjusted sequence ratios (ASR) with 95% confidence intervals were calculated. Analyses were stratified by sex, age (18-65 and 65+), proton pump inhibitor use and different initiation windows (90 and 365 days).

**Results:** There were 19,383 patients with an NSAID prescription and CV or VTE event (median age 66 [IQR 54 - 76] years, 54% male). Naproxen showed positive signals across all CV/VTE events, with highest ASR shown for PE (ASR 3.03 [95% CI 2.63-3.51]). Ibuprofen showed signals across six events, with PE the highest (ASR 2.2 [1.88-2.59]). Diclofenac and etoricoxib showed positive signals for five events with MI (ASR 3.30 [2.42-4.57]) and stroke (3.68 [2.14-6.62]) being the highest. Celecoxib and meloxicam showed positive signals across four events, with heart failure (2.15 [1.17-4.11]) and PE (2.66 [1.30-5.82]) the highest. Stratification analysis mostly aligned with the main analysis.

**Conclusions:** Individual NSAIDs showed variable CV and VTE signals, likely reflecting prescribing patterns and use. These findings suggest class-wide CV/VTE risks underscoring careful individual assessment when initiating therapy.

**Structured Graphical Abstract:** *Key Question:* Do individual NSAIDs show safety signals for cardiovascular or venous thromboembolic events in real-world data?

*Key Finding:* Initiation of several NSAIDs showed temporal safety signals for cardiovascular and venous thromboembolic events, with positive signals observed across multiple individual NSAIDs.

*Take-home Message:* Clinicians should balance the risks and benefits of individual NSAIDs, supporting a more personalised approach to prescribing and highlighting the need for further evaluation of identified safety signals.

## Introduction

Non-steroidal anti-inflammatory drugs (NSAIDs) are among the most widely prescribed medications, commonly used for antipyretic, anti-inflammatory, and analgesic purposes. Available in both over the counter and prescription forms, they are prescribed to alleviate a wide range of conditions, from headaches to chronic arthritis, and make up 5-10% of medication prescriptions worldwide(1).

Despite their widespread use, NSAIDs are associated with various adverse drug events (ADEs) including those related to gastrointestinal, cardiovascular (CV), and renal systems(2). These risks were highlighted by the withdrawal of rofecoxib in 2004 due to increased risk of CV events(3,4). Since then, numerous studies have shown NSAIDs are associated with increased CV risk(5). In addition, because arterial and venous thromboses share many underlying pathophysiological mechanisms, evidence also suggests NSAIDs may increase the risk of venous thromboembolism (VTE) events(6). Although both CV and VTE involve activation of coagulation pathways and endothelial dysfunction, they are distinct entities with different aetiologies and should be evaluated separately.

ADEs associated with NSAID use present a significant public health burden on healthcare systems, especially when prescribed to high-risk individuals, such as those older than 65 years of age(7). This has been estimated to cost the National Health Service (NHS) 31 million pounds and importantly contributed to a loss of 6,000 quality-adjusted life years for patients(2).

Evaluating the CV/VTE safety of individual NSAIDs using real-world data (RWD) is essential as randomised controlled trials (RCTs) often have limited generalisability, enrolling selected populations that may not reflect the diversity of patients in routine clinical practice(8). While previous literature has extensively examined CV events associated with NSAID use, associations with VTE have not been thoroughly investigated. Given that CV/VTE risk may differ between individual NSAIDs, an in-depth analysis of important drug-specific safety signals using RWD is needed to support post-marketing pharmacovigilance and evidence-based prescribing. This study aims to identify potential CV/VTE safety signals related to individual NSAIDs prescriptions using primary care data in the United Kingdom (UK).

## Methods

### Study design, setting and data sources

We performed a sequence symmetry analysis (SSA) with a case-only study design. People with a prescription for NSAIDs were identified from the Clinical Practice Research Datalink (CPRD) GOLD database(9) which includes primary care data from general practices across the UK representing ∼4.3% of the UK population(10). CPRD GOLD contains pseudonymised patient-level information on demographics, lifestyle data, clinical diagnoses, prescriptions, test results and preventive care. CPRD GOLD data were mapped to the Observational Medical Outcomes Partnership Common Data Model (OMOP CDM) version 5.4.1(11).

### Study participants

Individuals were required to be 18 years or older with at least one year of prior history. The study period was from January 2013 to January 2023. Individuals were included if they had initiated an NSAID and had a CV/VTE event of interest within ±180 days of NSAID initiation. Those with an NSAID prescription on the same day as a CV/VTE event of interest were excluded.

### Index exposure and marker outcome definitions

In this study, the index exposures were oral NSAIDs with at least 1,000 patient records during the study period and restricted to those with one single active ingredient. A full list of NSAIDs can be found in the supplement (**Table S1**).

The marker outcomes were CV events including myocardial infarction (MI), arrhythmia, heart failure, and stroke (including ischemic and haemorrhagic stroke); and VTE events including deep vein thrombosis (DVT) and pulmonary embolism (PE). The clinical code lists for these markers were created for previous regulatory work conducted with the European Medicines Agency (EMA)(12) using a standardised, reproducible framework for reliable and traceable phenotype generation(13). Codelists are provided in the supplement (**Table S2**).

### Statistical analysis Population characteristics

The population baseline characteristics of patients with a prescription for an NSAID and CV/VTE event of interest were summarised on a range of predefined comorbid conditions and medications, with median and interquartile range (IQR) used for continuous variables and counts and percentages used for categorical variables.

### Primary analysis

We used the R package “CohortSymmetry”(14) to perform SSA to identify potential CV/VTE safety signals associated with NSAID initiation. SSA is a signal detection method which evaluates asymmetry in the temporal sequence between drugs and adverse events by examining the order in which initiation of a drug (index exposure) and an event of interest (marker outcome) occurs(15). We applied a 365-day washout window for both the index and marker cohorts.

For each index-marker pair, we calculated the crude sequence ratio (CSR) by dividing the number of patients who developed the marker (CV/VTE event) after the index (NSAID prescription) by the number of patients who developed the marker before the index date. The adjusted sequence ratio (ASR) was calculated by adjusting the CSR by temporal prescribing trends using the null sequence ratio(16). An index-marker pair was considered a positive signal if the lower bound of the 95% CI for the ASR was greater than one.

Analysis was performed for each individual NSAID, all NSAIDs as a single group, and for non-selective NSAIDs and COX-2 inhibitors separately. We excluded aspirin from these groupings due to its possible use as a preventative treatment for CV(17).

### Sensitivity and subgroup analysis

Sensitivity analyses included testing additional NSAID initiation windows of 90 and 365 days. Subgroup analyses were stratified by sex, age (18–64 years and ≥65 years), and prior use of proton pump inhibitors (PPIs).

### Positive and negative controls

To evaluate the performance of SSA we first tested predefined positive and negative control drug pairs. We included study-specific controls to detect established study NSAID–events relationships including vomiting, nausea, oedema, anaemia, and acute kidney injury as positive controls, whereas cataracts were a negative control. Gastrointestinal (GI) haemorrhage was also included as a positive control for non-selective NSAIDs and as negative control for COX-2 inhibitors due to known differential effects across these NSAIDs(18). Finally, acetaminophen was included as a negative control for the CV/VTE study, as it is a commonly prescribed analgesic with similar indications to NSAIDs and has been used as a negative control in a recent European Medical Agency (EMA) commissioned study on the risk of VTE(19).

All analyses were performed using R version 4.2.3 with the analytical code available on GitHub to enable research reproducibility (https://github.com/oxford-pharmacoepi/SSA_nsaids_aesi).

## Results

All results for this study can be viewed on a user-friendly interactive app (dpa-pde-oxford.shinyapps.io/ssa_nsaids_cvd_study/).

### Study population and characteristics

A total of 19,383 individuals were identified (**Table 1**). The median age was 66 [54 - 76] years with the largest proportion of participants in the 70-79 age group (24.50%) and a slightly higher proportion of males (54.0%). Hypertension was the most common condition (27.8%) recorded any time prior to index event, followed by depressive disorder (17.8%), urinary tract infections (13.3%), and chronic kidney disease (13.1%). In the year prior, antibacterials (48.8%), drugs for acid related disorders (44.3%), opioids (40.0%), and lipid modifying agents (36.4%) were the most common medications.

**Table 1:** Baseline characteristics of individuals with record of any NSAID prescription and cardiovascular events of interest (excluding aspirin).

| Description | Any NSAID |
| --- | --- |
| Number subjects | 19,383 |
| Age (Median [Q25 - Q75]) | 66 [54 - 76] |
| **Age group (N %)** |  |
| 18 to 49 | 3,417 (17.6%) |
| 50 to 59 | 3,398 (17.5%) |
| 60 to 69 | 4,421 (22.8%) |
| 70 to 79 | 4,749 (24.5%) |
| 80 to 89 | 2,874 (14.8%) |
| 90+ | 524 (2.70%) |
| **Sex (N %)** |  |
| Male | 10,471 (54.0%) |
| **Conditions any time prior to index date (N %)** |  |
| Hypertension | 5,392 (27.8%) |
| Cancer excl. skin cancer | 2,364 (12.2%) |
| Hyperlipidaemia | 2,044 (10.6%) |
| UTI | 2,581 (13.3%) |
| COPD | 1,420 (7.33%) |
| Type 2 diabetes | 2,133 (11.0%) |
| Anaemia | 1,605 (8.28%) |
| Peripheral vascular disease | 374 (1.93%) |
| Cerebrovascular disease | 1,059 (5.46%) |
| Chronic kidney disease | 2,540 (13.1%) |
| Asthma | 2,192 (11.3%) |
| Ischemic heart disease | 1,978 (10.2%) |
| Depressive disorder | 3,451 (17.8%) |
| Obesity | 2,187 (11.3%) |
| GERD | 759 (3.92%) |
| **Medications 365 days prior to index date (N %)** |  |
| Systematic Antibacterials | 9,459 (48.8%) |
| Agents for the renin ang system | 6,579 (33.9%) |
| Antiepileptics | 2,066 (10.7%) |
| Lipid modifying agents | 7,059 (36.4%) |
| Drugs for addictive disorders | 567 (2.93%) |
| Antibiotics | 1,444 (7.45%) |
| Antiaenemic preparations | 3,523 (18.2%) |
| Immunosuppressants | 354 (1.83%) |
| Antidepressants | 5,477 (28.3%) |
| Drugs obstructive airway disorders | 5,294 (27.3%) |
| Drugs for acid related disorders | 8,579 (44.3%) |
| Psycholeptics | 3,890 (20.1%) |
| Drugs used in diabetes | 2,233 (11.5%) |
| Calcium channel blockers | 4,278 (22.1%) |
| Antithrombotics | 2,490 (12.9%) |
| Hormonal contraceptives | 518 (2.67%) |
| Diuretics | 4,628 (23.9%) |
| Beta blocking agents | 4,596 (23.7%) |
| Opioids | 7,744 (40.0%) |

The most prescribed NSAID excluding aspirin was naproxen (n = 9,773) followed by ibuprofen (n = 5,980) and diclofenac (n = 1,718) (**Table S3**). Individual characteristics for specific NSAIDs showed similar results with some exceptions. Diclofenac, etodolac, and mefenamate users were younger with higher proportions of those aged 60-69 years and 18-49 years (for mefenamate), respectively. Celecoxib and mefenamate had higher proportions of females (57.5% and 93.8%) whereas indomethacin users were predominantly male (71.3%). Regarding comorbidities and prior medication, indomethacin and mefenamate had slightly different characteristics compared to other NSAIDs. Indomethacin users had higher prescriptions for beta blockers (35.7%) and diuretics (39.7%), whereas among mefenamate users, depression was the most common comorbidity (24.2%), with antidepressants (33.2%) and drugs for obstructive airway disorders (24.7%) the most frequently prescribed medications.

Results of the positive and negative control analyses used to assess the validity of SSA are presented in **Table 2** and aligned with expectations. Results for the analgesic negative control, acetaminophen, showed negative signals across all CV/VTE events (**Table S4**).

**Table 2:** Results of predefined positive and negative control analyses used to evaluate performance of the sequence symmetry analysis.

| Index-marker pair | Control | Index N (%) | Marker N (%) | ASR [95% CI] |
| --- | --- | --- | --- | --- |
| Any NSAID - Vomiting | Positive | 6,483 (56.2%) | 5,053 (43.8%) | 1.30 [1.25 - 1.35] |
| Any NSAID - Nausea | Positive | 5,736 (57.9%) | 4,177 (42.1%) | 1.39 [1.33 - 1.44] |
| Any NSAID - Oedema | Positive | 5,599 (56.0%) | 4,401 (44.0%) | 1.25 [1.20 - 1.30] |
| Any NSAID - Anaemia | Positive | 5,671 (59.7%) | 3,834 (40.3%) | 1.43 [1.37 - 1.49] |
| Any NSAID - Acute kidney injury | Positive | 1,391 (65.0%) | 748 (35.0%) | 1.73 [1.59 - 1.90] |
| Any NSAID - Cataracts | Negative | 2,338 (51.0%) | 2,242 (49.0%) | 0.99 [0.93 - 1.05] |
| Non-selective - GI Haemorrhage | Positive | 6,333 (58.8%) | 4,445 (41.2%) | 1.22 [1.17 - 1.27] |
| COX-2 - GI Haemorrhage | Negative | 268 (54.4%) | 225 (45.6%) | 1.11 [0.93 - 1.32] |

3 Any NSAID includes all NSAIDs excluding aspirin, ASR adjusted sequence ratio. 4 Positive controls represent drug–event pairs with known or expected temporal 5 associations; negative controls represent pairs with no known temporal relationship. GI; 6 gastrointestinal.

Positive signals were observed across all CV events for diclofenac, ibuprofen, celecoxib, etoricoxib, and naproxen (**Figure 1**). The highest ASR was observed for MI with diclofenac (3.30; 95% CI [2.42–4.57]), ibuprofen (1.92 [1.65–2.25]), and naproxen (2.34 [2.08–2.64]). Arrhythmia and stroke demonstrated the greatest number of positive signals across individual NSAIDs. For arrhythmia, ASRs ranged from 1.34 [1.04–1.74] for etoricoxib to 1.90 [1.41–2.58] for meloxicam whereas for stroke, ASRs ranged from Subgroup analysis by stroke subtype (ischemic and haemorrhagic) showed that ischemic stroke signals were consistent with overall stroke findings apart from celecoxib and etodolac which showed null results. For haemorrhagic stroke, most signals were null apart from aspirin and ibuprofen which were both negative with ASR of 0.45 [0.38–0.53] and 0.65 [0.51–0.84] respectively (**Figure S1**).

**Figure 1.**
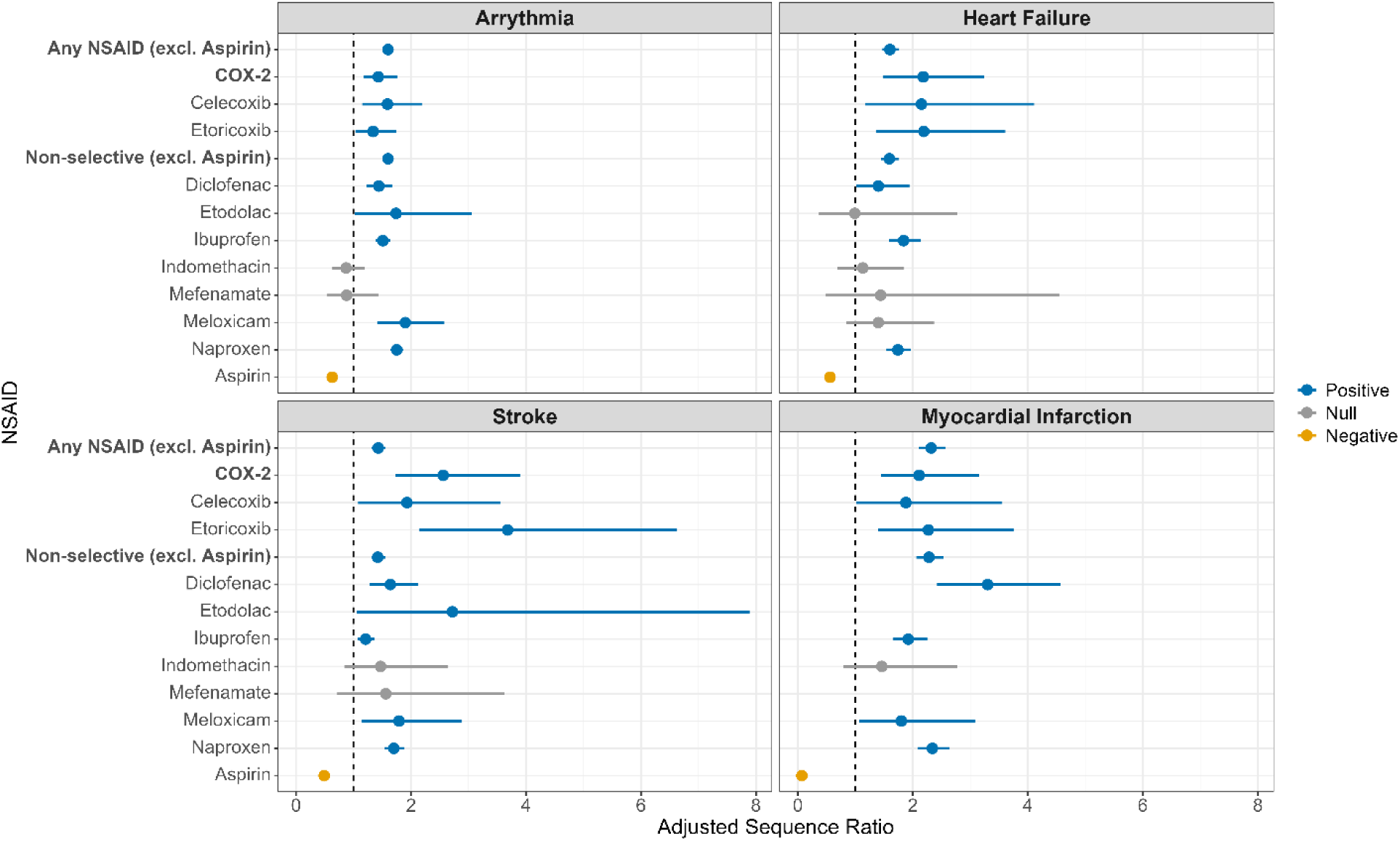
Adjusted sequence ratios with 95% confidence intervals for cardiovascular events. Estimates are shown for individual NSAIDs, any NSAID, non-selective NSAIDs and selective COX-2 inhibitors across arrhythmia, heart failure, stroke, and myocardial infarction.

In relation to VTE outcomes, diclofenac, ibuprofen, and naproxen showed positive signals for both DVT and PE with the highest signals seen for PE for all three drugs with ASR of 1.99 [1.53–2.60], 2.20 [1.88–2.59], and 3.03 [2.63–3.51], respectively (**Figure 2**). Signals were only observed for DVT for celecoxib (1.93 [1.14 - 3.34]) and PE for etoricoxib (2.69 [1.47–5.15]) and meloxicam (2.66 [1.30–5.82]). Indomethacin and mefenamate showed no signals for any VTE events and aspirin showed a negative signal for DVT and a null signal for PE.

**Figure 2.**
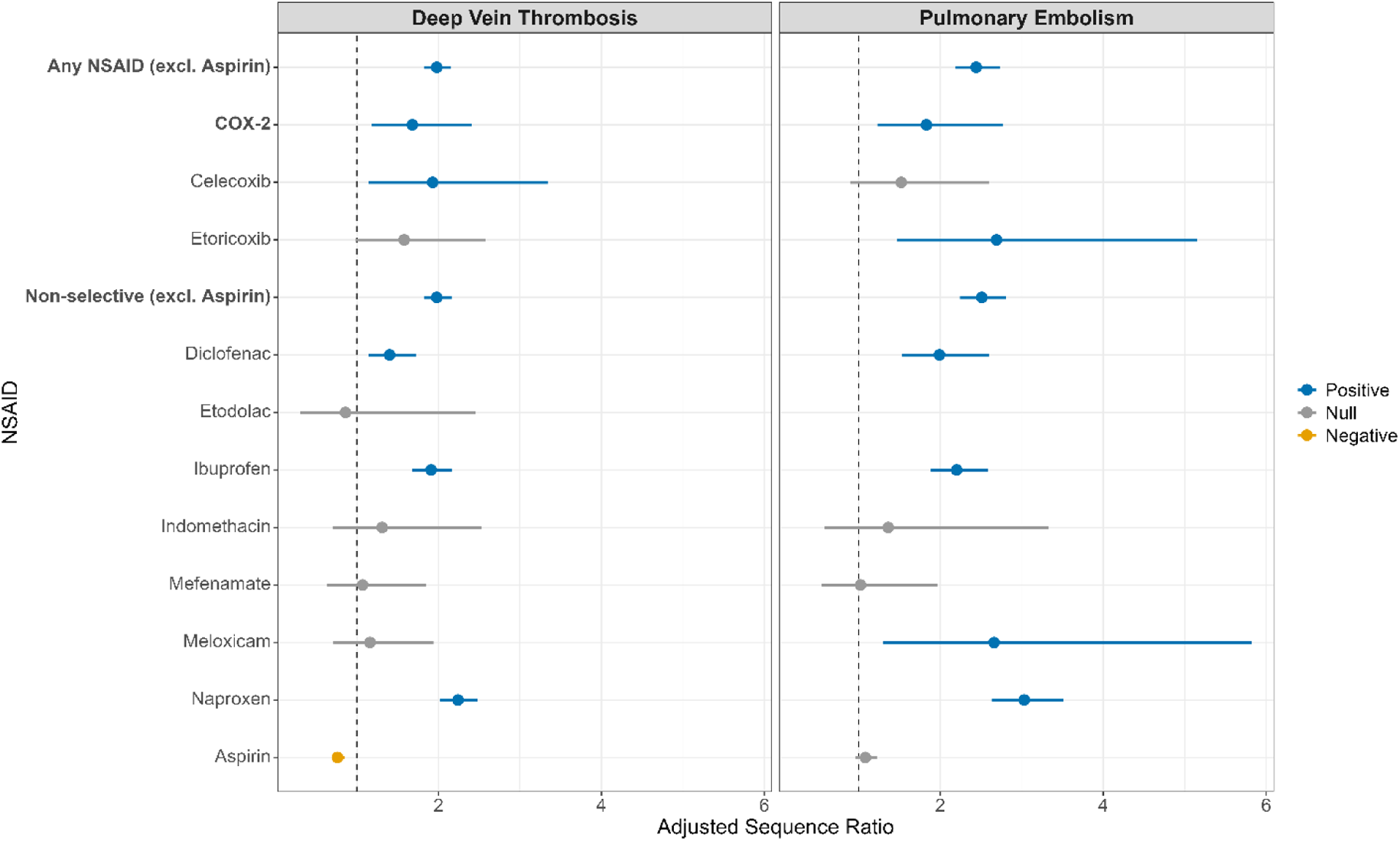
Adjusted sequence ratios with 95% confidence intervals for venous thromboembolism events. Estimates are shown for individual NSAIDs, any NSAID, non-selective NSAIDs and selective COX-2 inhibitors across pulmonary embolism and deep vein thrombosis.

Grouping NSAIDs together into any NSAID and further stratifying by COX-2 inhibitors and non-selective NSAIDs showed positive signals observed across all CV/VTE events. All ASRs, counts and percentages of each NSAID and CV/VTE event presented in **Figures 1 and 2** can be found in the supplement (**Table S5**).

Stratification by sex and age showed broadly consistent positive signals, with only a few exceptions (**Figures S2–S3**). Positive signals were observed for any NSAID, non-selective NSAIDs, and naproxen across all subgroups. Among selective COX-2 inhibitors, females showed no positive signals for DVT or PE, and males showed none for arrhythmia. Celecoxib showed a positive signal for arrhythmia in females (ASR 2.09 [1.36–3.27]) but not for other events, whereas in males, signals were seen for DVT (2.43 [1.05–6.05]), stroke (2.54 [1.11–6.29]), and MI (2.41 [1.05–6.02]). Etoricoxib showed signals for heart failure (3.15 [1.49–7.22]) and MI (3.87 [1.56–10.99]) in females, and for DVT (2.68 [1.19–6.58]) in males. Diclofenac showed positive signals except DVT in females and heart failure in males, which were null. Ibuprofen also showed positive signals except for stroke in males with meloxicam also showing signals for stroke (1.99 [1.01–4.13]) and MI (2.44 [1.18–5.38]) in males only. Indomethacin showed a positive signal for stroke in females only. Aspirin remained negative except for PE in females (1.22 [1.03–1.46]).

By age, patients ≥65 years showed positive signals for all CV and VTE events for selective COX-2 inhibitors, whereas those <65 years had signals for DVT, heart failure, and stroke only. Celecoxib was positive for arrhythmia in younger patients (2.18 [1.27–3.85]); etoricoxib for arrhythmia (1.58 [1.15–2.17]) and MI (2.30 [1.29–4.25]) in older adults. Ibuprofen showed positive signals across all events for both age strata except arrhythmia and stroke in younger adults. Meloxicam showed signals for arrhythmia (2.17 [1.52–3.14]) and MI (1.95 [1.01–3.92]) in older adults, and for stroke (2.49 [1.04–6.61]) in younger adults.

Sensitivity analysis varying the NSAID initiation window to either 90 days or 365 days showed also consistent positive signals with some exceptions. Results for the 90-day initiation window showed fewer positive signals (**Figure S4**) whereas using a 365-day initiation window showed some additional positive signals for some NSAIDs (**Figure S5**). For example, mefenamate and meloxicam showed positive signals across most CV/VTE events and indomethacin showed positive signals for stroke and MI. Subgroup analysis based on prior PPI use showed that positive signals for any NSAID were present in both the PPI and non-PPI groups, consistent with the main analysis. However, ASRs were lower among those with prior PPI use (**Figure S6**).

## Discussion

CV and VTE safety signals did vary across individual NSAIDs, with most NSAIDs showing positive signals with one or more CV or VTE events within CPRD GOLD using SSA. These patterns were relatively consistent across sex, age groups, and initiation windows. Stratification by PPI use showed slight inflation of positive signals, likely attributable to reverse causation.

Previous studies using SSA have also reported positive signals between NSAID initiation and CV events(5). For example, an SSA study from Australia that used veteran claims data linked to prescriptions and stroke hospitalisations found positive signals for overall, ischaemic, and haemorrhagic stroke across both non-selective and COX-2 selective NSAIDs, except for ibuprofen which showed null results (20). In contrast, indomethacin showed a positive signal in that study but null in ours. Our findings are broadly consistent with this prior work and extend it by including a wider range of CV outcomes such as arrhythmia and the more understudied VTE events. We also provided stratified analyses by age and sex. Given the evolving prescribing patterns and safety concerns of NSAIDs, it is important to evaluate safety signals in a more recent study period(21). The difference in signals for ibuprofen and indomethacin between studies may reflect variation in prescribing behaviour, initiation windows, age structure, as well as methodological differences in event capture.

To date, there are no published studies that have applied SSA to investigate associations between NSAID initiation and VTE events. However, evidence from other observational study designs suggested an increased risk of VTE following NSAID use(6). This included a nationwide cohort study in Denmark which reported that NSAID use and concomitant use of NSAIDs and hormone contraceptives was associated with an elevated VTE risk in women of reproductive age compared to no NSAID use(22). More recently, an EMA-commissioned study across four European countries using a self-controlled case series design also identified a positive association between VTE and NSAID exposure among hormonal contraceptive users. However, it also found a counterintuitive increase in VTE risk in the two weeks prior to NSAID initiation, suggesting that protopathic bias and residual confounding may partly explain the association(19).

Positive signals across the various NSAIDs suggest that the CV/VTE risk of NSAIDs may extend across the whole therapeutic class of NSAIDs(23). Various studies have demonstrated that all NSAIDs are associated with CV/VTE events with evidence from RCTs and observational studies indicating that while certain NSAIDs may have lower CV/VTE risk profiles none are completely without CV/VTE risk(5,6,24,25).

Although some NSAIDs, such as indomethacin and mefenamate, showed no CV/VTE signals, this could reflect their short-term and episodic usage. This intermittent use could result in insufficient exposure to detect an increase in CV/VTE events soon after NSAID initiation. Despite this, sensitivity analyses using a longer initiation window showed positive signals for stroke and MI for indomethacin, potentially suggesting that null findings may reflect delayed pathophysiological processes leading to CV/VTE events after NSAID initiation. This interpretation aligns with evidence suggesting CV risksfor both these NSAIDs (32,33). However, care must be taken with interpretation of results with longer initiation windows, as extending the window can increase the potential for time-varying confounding, which might bias results and highlights the need to align initiation windows with the underlying disease processes(28).

The negative signal observed for aspirin appears to reflect current prescribing practices, where it is used preventatively due to its antiplatelet effects(29). Given its role in secondary prevention, and declining use in primary prevention or other chronic conditions, the negative signals may also be influenced by the non-mutually exclusive CV events within this study, as well as its therapeutic role post-events. Acetaminophen was included as an additional negative control because of its wide use for pain management but does not share anti-inflammatory mechanisms associated with increased CV/VTE risk observed for NSAIDs(30,31). The absence of positive signals for both aspirin and acetaminophen supports the validity of the analytical approach and suggests that the observed associations for other NSAIDs reflect true safety signals. Stratification by sex and age showed consistent positive signals with the main analysis, indicating that the temporal asymmetry between NSAID initiation and CV/VTE events was not only confined to specific demographic groups. However, positive signals for COX-2 inhibitors were observed across all CV and VTE event types among individuals aged ≥65 years, underscoring the need for cautious prescribing and monitoring in older, high-risk populations(32). Additionally, some variation was observed across strata for certain NSAIDs, which may reflect differences in frequency of events, comorbidity burden, or cumulative exposure rather than true effect modification which is beyond the scope of the SSA method. Although age- and sex-related differences in CV/VTE risk are well recognised(33,34), such differences were not apparent in the temporal associations between NSAID initiation and CV/VTE events in this study. This likely reflects the multifactorial nature of NSAID-related CV/VTE effects, which are influenced by baseline CV/VTE risk, comorbidities, exposure duration, and concurrent medication use rather than a single pharmacological biological mechanism. These factors may obscure or attenuate subgroup variation in temporal asymmetry. Given SSA assesses within-person temporal asymmetry rather than risk, these findings should therefore be interpreted as supporting the robustness of the overall signal pattern rather than indicating demographic differences in baseline risk or the absence of sex-specific biological effects.

Strengths of this study include the use of a large, comprehensive primary care database that enabled real-world signal detection across a large UK population. The use of SSA reduces bias from time-invariant confounders and accounts for changes in prescribing trends, allowing the identification of potential temporal associations between NSAID initiation and CV/VTE events. However, our study does have limitations. Firstly, SSA is a signal detection method used to identify temporal asymmetry therefore cannot establish causality with further validation of positive signals required using more robust epidemiological designs such as self-controlled case series(35). Over-the-counter NSAID use was not captured, which may have led to exposure misclassification and potential attenuation of signals. Only oral NSAIDs were included, and results may vary by NSAID dose and exposure duration, which could potentially influence CV/VTE event risk(36). Some of the CV/VTE events may have longer latency or development periods rather than being acute, potentially introducing time-varying confounding. Subgroup analysis stratified by prior PPI use showed attenuation of signals among prior users, suggesting possible reverse causation but did not change our conclusions.

## Conclusion

In summary, this study identified temporal asymmetry between NSAID initiation and several CV and VTE events, with variability across individual NSAIDs likely influenced by differences in prescribing behaviours, duration of use, and underlying indications. These findings suggest that CV/VTE risks may extend across the NSAID class and underscore the need for careful individual assessment of CV/VTE risk when initiating therapy, particularly given their widespread and often over-the-counter use. Further studies using complementary epidemiologic designs are needed to confirm and validate these signal detection findings.

## Abbreviations

CV: cardiovascular
VTE: venous thromboembolic
SSA: sequence symmetry analysis
ASR: adjusted sequence ratio
NSAIDs: Non-steroidal anti-inflammatory drugs
MI: myocardial infarction
DVT: deep vein thrombosis (DVT)
PE: pulmonary embolism

## Funding

SSB was funded by the University of Alabama at Birmingham Marnix E. Heersink School of Medicine. Additionally, there was partial support from the Oxford NIHR Biomedical Research Centre.

## Acknowledgements

None.

## Author Contributions

Conceptualization; DN, DPA; Data harmonisation and data quality assessment; AD, WYM; Formal analysis; ER, SSB; Funding acquisition; DPA, SSB; Supervision; DPA, DN; Interpretation of results: All authors; Roles/Writing - original draft: SSB, ER, DN; Writing - review & editing: All authors

## Conflict of Interest Statement

Professor Daniel Prieto-Alhambra’s research group from the University of Oxford has received research grants from the European Medicines Agency, from the Innovative Medicines Initiative, from Gilead Science, from Theramex and from UCB Biopharma. All other authors declare no conflicts of interest.

## Sponsor Role

The sponsors of the study did not have any involvement in the writing of the manuscript or the decision to submit it for publication.

## Data availability statement

This study is based in part on data from the Clinical Practice Research Datalink (CPRD) obtained under licence from the UK Medicines and Healthcare products Regulatory Agency. The data are provided by patients and collected by the NHS as part of their care and support. The interpretation and conclusions contained in this study are those of the author/s alone. Patient level data used in this study were obtained through the University of Oxford multi-study licence and an approved CPRD application (protocol number 25_004835) following the CPRD Research Data Governance process. Details on how to apply for data access can be found at https://cprd.com/data-access.

Individual patient consent was not required, as CPRD data are de-identified and provided under ethical approval from the UK Health Research Authority (HRA) and the NHS Health and Social Care Research Ethics Committee.

## Notes

### Author Declarations

Individual patient consent was not required, as CPRD data are de-identified and provided under ethical approval from the UK Health Research Authority (HRA) and the NHS Health and Social Care Research Ethics Committee. Patient level data used in this study were obtained through the University of Oxford multi-study licence and an approved CPRD application (protocol number 25_004835) following the CPRD Research Data Governance process.

