## Supplement for "Cardiovascular and Thromboembolic Safety Signals Associated with Non-Steroidal Anti-Inflammatory Drugs Initiation: A Sequence Symmetry Analysis"

### Table S1: Initial list of NSAIDs (excluding aspirin) used in this study with more than 1000 patient records

**NSAID**

celecoxib

etoricoxib

diclofenac

etodolac

ibuprofen

indomethacin

mefenamate

meloxicam

naproxen

piroxicam

aceclofenac

dexibuprofen

dexketoprofen

diflunisal

flurbiprofen

ketoprofen

nabumetone

sulindac

tenoxicam

tiaprofenate

tolfenamic acid

### Table S2: Codelists of cardiovascular, thromboembolic and gastrointestinal hemorrhage events used in this study

| **Concept ID** | **Concept Code** | **Name** | **Vocabulary** | **Condition** |
| --- | --- | --- | --- | --- |
| 45766075 | 703164000 | Acute anterior ST segment elevation myocardial infarction | SNOMED | Myocardial infarction (MI) |
| 4178129 | 52035003 | Acute anteroapical myocardial infarction | SNOMED | Myocardial infarction (MI) |
| 4267568 | 62695002 | Acute anteroseptal myocardial infarction | SNOMED | Myocardial infarction (MI) |
| 312327 | 57054005 | Acute myocardial infarction | SNOMED | Myocardial infarction (MI) |
| 44782769 | 17531000119105 | Acute myocardial infarction due to left coronary artery occlusion | SNOMED | Myocardial infarction (MI) |
| 44782712 | 23311000119105 | Acute myocardial infarction due to right coronary artery occlusion | SNOMED | Myocardial infarction (MI) |
| 45766115 | 703212004 | Acute myocardial infarction during procedure | SNOMED | Myocardial infarction (MI) |
| 434376 | 54329005 | Acute myocardial infarction of anterior wall | SNOMED | Myocardial infarction (MI) |
| 45766150 | 703252002 | Acute myocardial infarction of anterior wall involving right ventricle | SNOMED | Myocardial infarction (MI) |
| 438438 | 70211005 | Acute myocardial infarction of anterolateral wall | SNOMED | Myocardial infarction (MI) |
| 4243372 | 59063002 | Acute myocardial infarction of apical-lateral wall | SNOMED | Myocardial infarction (MI) |
| 4108669 | 194809007 | Acute myocardial infarction of atrium | SNOMED | Myocardial infarction (MI) |
| 4275436 | 64627002 | Acute myocardial infarction of high lateral wall | SNOMED | Myocardial infarction (MI) |
| 438170 | 73795002 | Acute myocardial infarction of inferior wall | SNOMED | Myocardial infarction (MI) |
| 45771322 | 703251009 | Acute myocardial infarction of inferior wall involving right ventricle | SNOMED | Myocardial infarction (MI) |
| 438447 | 65547006 | Acute myocardial infarction of inferolateral wall | SNOMED | Myocardial infarction (MI) |
| 441579 | 76593002 | Acute myocardial infarction of inferoposterior wall | SNOMED | Myocardial infarction (MI) |
| 436706 | 58612006 | Acute myocardial infarction of lateral wall | SNOMED | Myocardial infarction (MI) |
| 4324413 | 70998009 | Acute myocardial infarction of posterobasal wall | SNOMED | Myocardial infarction (MI) |
| 4051874 | 15990001 | Acute myocardial infarction of posterolateral wall | SNOMED | Myocardial infarction (MI) |
| 4303359 | 79009004 | Acute myocardial infarction of septum | SNOMED | Myocardial infarction (MI) |
| 4145721 | 307140009 | Acute non-Q wave infarction | SNOMED | Myocardial infarction (MI) |
| 4119944 | 233828006 | Acute non-Q wave infarction - anterolateral | SNOMED | Myocardial infarction (MI) |
| 4119456 | 233826005 | Acute non-Q wave infarction - anteroseptal | SNOMED | Myocardial infarction (MI) |
| 4119945 | 233830008 | Acute non-Q wave infarction - inferior | SNOMED | Myocardial infarction (MI) |
| 4119946 | 233832000 | Acute non-Q wave infarction - inferolateral | SNOMED | Myocardial infarction (MI) |
| 4121466 | 233834004 | Acute non-Q wave infarction - lateral | SNOMED | Myocardial infarction (MI) |
| 4124685 | 233837006 | Acute non-Q wave infarction - widespread | SNOMED | Myocardial infarction (MI) |
| 4270024 | 401314000 | Acute non-ST segment elevation myocardial infarction | SNOMED | Myocardial infarction (MI) |
| 35610091 | 1089451000000100 | Acute nontransmural myocardial infarction | SNOMED | Myocardial infarction (MI) |
| 319039 | 233838001 | Acute posterior myocardial infarction | SNOMED | Myocardial infarction (MI) |
| 4119457 | 233827001 | Acute Q wave infarction - anterolateral | SNOMED | Myocardial infarction (MI) |
| 4119943 | 233825009 | Acute Q wave infarction - anteroseptal | SNOMED | Myocardial infarction (MI) |
| 4121464 | 233829003 | Acute Q wave infarction - inferior | SNOMED | Myocardial infarction (MI) |
| 4121465 | 233831007 | Acute Q wave infarction - inferolateral | SNOMED | Myocardial infarction (MI) |
| 4124684 | 233833005 | Acute Q wave infarction - lateral | SNOMED | Myocardial infarction (MI) |
| 4119948 | 233836002 | Acute Q wave infarction - widespread | SNOMED | Myocardial infarction (MI) |
| 4126801 | 304914007 | Acute Q wave myocardial infarction | SNOMED | Myocardial infarction (MI) |
| 4296653 | 401303003 | Acute ST segment elevation myocardial infarction | SNOMED | Myocardial infarction (MI) |
| 46270162 | 15713081000119100 | Acute ST segment elevation myocardial infarction due to left coronary artery occlusion | SNOMED | Myocardial infarction (MI) |
| 46270163 | 15713121000119100 | Acute ST segment elevation myocardial infarction due to right coronary artery occlusion | SNOMED | Myocardial infarction (MI) |
| 43020460 | 285981000119103 | Acute ST segment elevation myocardial infarction involving left anterior descending coronary artery | SNOMED | Myocardial infarction (MI) |
| 45766076 | 703165004 | Acute ST segment elevation myocardial infarction of anterior wall involving right ventricle | SNOMED | Myocardial infarction (MI) |
| 761736 | 15962541000119100 | Acute ST segment elevation myocardial infarction of anteroapical wall | SNOMED | Myocardial infarction (MI) |
| 46270159 | 15712881000119100 | Acute ST segment elevation myocardial infarction of anterolateral wall | SNOMED | Myocardial infarction (MI) |
| 46270160 | 15712961000119100 | Acute ST segment elevation myocardial infarction of anteroseptal wall | SNOMED | Myocardial infarction (MI) |
| 45766116 | 703213009 | Acute ST segment elevation myocardial infarction of inferior wall | SNOMED | Myocardial infarction (MI) |
| 45766151 | 703253007 | Acute ST segment elevation myocardial infarction of inferior wall involving right ventricle | SNOMED | Myocardial infarction (MI) |
| 35611570 | 12238111000119100 | Acute ST segment elevation myocardial infarction of inferolateral wall | SNOMED | Myocardial infarction (MI) |
| 35611571 | 12238151000119100 | Acute ST segment elevation myocardial infarction of inferoposterior wall | SNOMED | Myocardial infarction (MI) |
| 46274044 | 15712921000119100 | Acute ST segment elevation myocardial infarction of lateral wall | SNOMED | Myocardial infarction (MI) |
| 46270161 | 15713041000119100 | Acute ST segment elevation myocardial infarction of posterior wall | SNOMED | Myocardial infarction (MI) |
| 46273495 | 15713201000119100 | Acute ST segment elevation myocardial infarction of posterobasal wall | SNOMED | Myocardial infarction (MI) |
| 46270158 | 15712841000119100 | Acute ST segment elevation myocardial infarction of posterolateral wall | SNOMED | Myocardial infarction (MI) |
| 46270164 | 15713161000119100 | Acute ST segment elevation myocardial infarction of septum | SNOMED | Myocardial infarction (MI) |
| 444406 | 70422006 | Acute subendocardial infarction | SNOMED | Myocardial infarction (MI) |
| 35610093 | 1089471000000100 | Acute transmural myocardial infarction | SNOMED | Myocardial infarction (MI) |
| 4119947 | 233835003 | Acute widespread myocardial infarction | SNOMED | Myocardial infarction (MI) |
| 37109912 | 723860000 | Arrhythmia due to and following acute myocardial infarction | SNOMED | Myocardial infarction (MI) |
| 438172 | 194863005 | Atrial septal defect due to and following acute myocardial infarction | SNOMED | Myocardial infarction (MI) |
| 4124687 | 233847009 | Cardiac rupture due to and following acute myocardial infarction | SNOMED | Myocardial infarction (MI) |
| 4215259 | 394710008 | First myocardial infarction | SNOMED | Myocardial infarction (MI) |
| 4108678 | 194862000 | Hemopericardium due to and following acute myocardial infarction | SNOMED | Myocardial infarction (MI) |
| 4173632 | 42531007 | Microinfarct of heart | SNOMED | Myocardial infarction (MI) |
| 45766214 | 703328007 | Mitral valve regurgitation due to acute myocardial infarction without papillary muscle and chordal rupture | SNOMED | Myocardial infarction (MI) |
| 45771327 | 703330009 | Mitral valve regurgitation due to acute myocardial infarction with papillary muscle and chordal rupture | SNOMED | Myocardial infarction (MI) |
| 4323202 | 428196007 | Mixed myocardial ischemia and infarction | SNOMED | Myocardial infarction (MI) |
| 4329847 | 22298006 | Myocardial infarction | SNOMED | Myocardial infarction (MI) |
| 37309626 | 16837681000119100 | Myocardial infarction due to demand ischemia | SNOMED | Myocardial infarction (MI) |
| 4170094 | 418044006 | Myocardial infarction in recovery phase | SNOMED | Myocardial infarction (MI) |
| 4200113 | 314207007 | Non-Q wave myocardial infarction | SNOMED | Myocardial infarction (MI) |
| 4030582 | 129574000 | Postoperative myocardial infarction | SNOMED | Myocardial infarction (MI) |
| 35610087 | 1089431000000100 | Postoperative nontransmural myocardial infarction | SNOMED | Myocardial infarction (MI) |
| 4206867 | 311796008 | Postoperative subendocardial myocardial infarction | SNOMED | Myocardial infarction (MI) |
| 35610089 | 1089441000000100 | Postoperative transmural myocardial infarction | SNOMED | Myocardial infarction (MI) |
| 4207921 | 311792005 | Postoperative transmural myocardial infarction of anterior wall | SNOMED | Myocardial infarction (MI) |
| 4209541 | 311793000 | Postoperative transmural myocardial infarction of inferior wall | SNOMED | Myocardial infarction (MI) |
| 37109911 | 723859005 | Pulmonary embolism due to and following acute myocardial infarction | SNOMED | Myocardial infarction (MI) |
| 4108679 | 194865003 | Rupture of cardiac wall without hemopericardium as current complication following acute myocardial infarction | SNOMED | Myocardial infarction (MI) |
| 4108219 | 194866002 | Rupture of chordae tendinae due to and following acute myocardial infarction | SNOMED | Myocardial infarction (MI) |
| 4147223 | 30277009 | Rupture of ventricle due to acute myocardial infarction | SNOMED | Myocardial infarction (MI) |
| 4124686 | 233843008 | Silent myocardial infarction | SNOMED | Myocardial infarction (MI) |
| 765132 | 380001000004106 | Subendocardial myocardial infarction | SNOMED | Myocardial infarction (MI) |
| 4108217 | 194856005 | Subsequent myocardial infarction | SNOMED | Myocardial infarction (MI) |
| 4108677 | 194857001 | Subsequent myocardial infarction of anterior wall | SNOMED | Myocardial infarction (MI) |
| 4108218 | 194858006 | Subsequent myocardial infarction of inferior wall | SNOMED | Myocardial infarction (MI) |
| 45766241 | 703360004 | Subsequent non-ST segment elevation myocardial infarction | SNOMED | Myocardial infarction (MI) |
| 45766114 | 703211006 | Subsequent ST segment elevation myocardial infarction | SNOMED | Myocardial infarction (MI) |
| 45766113 | 703210007 | Subsequent ST segment elevation myocardial infarction of anterior wall | SNOMED | Myocardial infarction (MI) |
| 45773170 | 703209002 | Subsequent ST segment elevation myocardial infarction of inferior wall | SNOMED | Myocardial infarction (MI) |
| 4108680 | 194868001 | Thrombosis of atrium, auricular appendage, and ventricle due to and following acute myocardial infarction | SNOMED | Myocardial infarction (MI) |
| 439693 | 194802003 | True posterior myocardial infarction | SNOMED | Myocardial infarction (MI) |
| 37109910 | 723858002 | Ventricular aneurysm due to and following acute myocardial infarction | SNOMED | Myocardial infarction (MI) |
| 4151046 | 282006 | Acute myocardial infarction of basal-lateral wall | SNOMED | Myocardial infarction (MI) |
| 45766212 | 703326006 | Mitral valve regurgitation due to and following acute myocardial infarction | SNOMED | Myocardial infarction (MI) |
| 761737 | 15963181000119100 | Acute ST segment elevation myocardial infarction due to occlusion of circumflex coronary artery | SNOMED | Myocardial infarction (MI) |
| 4154704 | 371068009 | Myocardial infarction with complication | SNOMED | Myocardial infarction (MI) |
| 4092011 | 251183004 | Aberrantly conducted complex | SNOMED | Arrythmia |
| 4091901 | 251167004 | Aberrant premature complexes | SNOMED | Arrythmia |
| 4057008 | 16797001 | Accelerated atrioventricular conduction | SNOMED | Arrythmia |
| 4247537 | 61277005 | Accelerated idioventricular rhythm | SNOMED | Arrythmia |
| 37312140 | 789693005 | Acquired Brugada syndrome | SNOMED | Arrythmia |
| 37116420 | 733125004 | Acquired complete atrioventricular block | SNOMED | Arrythmia |
| 40479264 | 442946007 | Acquired long QT syndrome | SNOMED | Arrythmia |
| 313224 | 17869006 | Anomalous atrioventricular excitation | SNOMED | Arrythmia |
| 4296729 | 76887001 | Anterior fascicular block, posterior fascicular block AND incomplete right bundle branch block | SNOMED | Arrythmia |
| 4121481 | 233899006 | Antidromic atrioventricular re-entrant tachycardia | SNOMED | Arrythmia |
| 37108582 | 10701000087104 | Arrhythmia during surgery | SNOMED | Arrythmia |
| 4102252 | 300997008 | Asymptomatic sinoatrial node dysfunction | SNOMED | Arrythmia |
| 4068155 | 17366009 | Atrial arrhythmia | SNOMED | Arrythmia |
| 4088504 | 251173003 | Atrial bigeminy | SNOMED | Arrythmia |
| 4088986 | 251187003 | Atrial escape complex | SNOMED | Arrythmia |
| 313217 | 49436004 | Atrial fibrillation | SNOMED | Arrythmia |
| 44782442 | 120041000119109 | Atrial fibrillation with rapid ventricular response | SNOMED | Arrythmia |
| 314665 | 5370000 | Atrial flutter | SNOMED | Arrythmia |
| 4088987 | 251188008 | Atrial parasystole | SNOMED | Arrythmia |
| 4108830 | 195069001 | Atrial paroxysmal tachycardia | SNOMED | Arrythmia |
| 4115173 | 287057009 | Atrial premature complex | SNOMED | Arrythmia |
| 42872924 | 450919004 | Atrial standstill | SNOMED | Arrythmia |
| 4171269 | 276796006 | Atrial tachycardia | SNOMED | Arrythmia |
| 4091903 | 251174009 | Atrial trigeminy | SNOMED | Arrythmia |
| 316135 | 233917008 | Atrioventricular block | SNOMED | Arrythmia |
| 43020929 | 472809000 | Atrioventricular block due to endocarditis | SNOMED | Arrythmia |
| 4305210 | 418341009 | Atrioventricular conduction disorder | SNOMED | Arrythmia |
| 4175473 | 50799005 | Atrioventricular dissociation | SNOMED | Arrythmia |
| 42536726 | 735685003 | Atrioventricular reciprocating tachycardia | SNOMED | Arrythmia |
| 4081675 | 278482008 | Atrioventricular tachycardia | SNOMED | Arrythmia |
| 36712986 | 15964901000119100 | Atypical atrial flutter | SNOMED | Arrythmia |
| 4088502 | 251163000 | AV junctional (nodal) arrest | SNOMED | Arrythmia |
| 4088983 | 251162005 | AV-junctional (nodal) bradycardia | SNOMED | Arrythmia |
| 4088503 | 251165007 | AV junctional (nodal) tachycardia | SNOMED | Arrythmia |
| 4038688 | 11849007 | AV junctional rhythm | SNOMED | Arrythmia |
| 4228836 | 88412007 | AV node arrhythmia | SNOMED | Arrythmia |
| 4250169 | 74021003 | Bifascicular block | SNOMED | Arrythmia |
| 40483798 | 445481009 | Bifascicular block on electrocardiogram | SNOMED | Arrythmia |
| 321587 | 20143001 | Bilateral bundle branch block | SNOMED | Arrythmia |
| 4088984 | 251170000 | Blocked premature atrial contraction | SNOMED | Arrythmia |
| 4228448 | 421869004 | Bradyarrhythmia | SNOMED | Arrythmia |
| 313791 | 6374002 | Bundle branch block | SNOMED | Arrythmia |
| 40484036 | 442559009 | Bundle branch reentrant ventricular tachycardia | SNOMED | Arrythmia |
| 4301015 | 78240009 | Cardiac arrest due to pacemaker failure | SNOMED | Arrythmia |
| 44784217 | 698247007 | Cardiac arrhythmia | SNOMED | Arrythmia |
| 45757098 | 10743741000119100 | Cardiac arrhythmia in mother complicating childbirth | SNOMED | Arrythmia |
| 4303238 | 419671004 | Catecholaminergic polymorphic ventricular tachycardia | SNOMED | Arrythmia |
| 42536725 | 735683005 | Cavotricuspid isthmus dependent macroreentry tachycardia | SNOMED | Arrythmia |
| 36715042 | 720507006 | Chronic atrial and intestinal dysrhythmia | SNOMED | Arrythmia |
| 4141360 | 426749004 | Chronic atrial fibrillation | SNOMED | Arrythmia |
| 4137382 | 425615007 | Chronic atrial flutter | SNOMED | Arrythmia |
| 4098133 | 26950008 | Chronic ectopic atrial tachycardia | SNOMED | Arrythmia |
| 320744 | 27885002 | Complete atrioventricular block | SNOMED | Arrythmia |
| 46269694 | 102451000119107 | Complete atrioventricular block as complication of atrioventricular nodal ablation | SNOMED | Arrythmia |
| 4267892 | 6180003 | Complete left bundle branch block | SNOMED | Arrythmia |
| 4088337 | 251123001 | Complete right bundle branch block | SNOMED | Arrythmia |
| 316999 | 44808001 | Conduction disorder of the heart | SNOMED | Arrythmia |
| 4117112 | 300996004 | Controlled atrial fibrillation | SNOMED | Arrythmia |
| 4304839 | 82226007 | Diffuse intraventricular block | SNOMED | Arrythmia |
| 4064452 | 164889003 | ECG: atrial fibrillation | SNOMED | Arrythmia |
| 4064867 | 164881000 | ECG: ectopic beats | SNOMED | Arrythmia |
| 4065290 | 164903001 | ECG: partial atrioventricular block - 2:1 | SNOMED | Arrythmia |
| 4064873 | 164902006 | ECG: partial atrioventricular block - long PR | SNOMED | Arrythmia |
| 4141820 | 427393009 | ECG: sinus arrhythmia | SNOMED | Arrythmia |
| 4064612 | 164884008 | ECG: ventricular ectopics | SNOMED | Arrythmia |
| 4236004 | 406461004 | Ectopic atrial beats | SNOMED | Arrythmia |
| 4121479 | 233892002 | Ectopic atrial tachycardia | SNOMED | Arrythmia |
| 4121480 | 233895000 | Ectopic atrioventricular node tachycardia | SNOMED | Arrythmia |
| 4143042 | 33413000 | Ectopic beats | SNOMED | Arrythmia |
| 4164083 | 29320008 | Ectopic rhythm | SNOMED | Arrythmia |
| 4065285 | 164885009 | EKG: atrial ectopics | SNOMED | Arrythmia |
| 4065288 | 164890007 | EKG: atrial flutter | SNOMED | Arrythmia |
| 4064874 | 164906009 | EKG: complete atrioventricular block | SNOMED | Arrythmia |
| 4064457 | 164898000 | EKG: heart block | SNOMED | Arrythmia |
| 4064614 | 164909002 | EKG: left bundle branch block | SNOMED | Arrythmia |
| 4138921 | 426183003 | EKG: Mobitz type II atrioventricular block | SNOMED | Arrythmia |
| 4064460 | 164907000 | EKG: right bundle branch block | SNOMED | Arrythmia |
| 4065287 | 164887001 | EKG: supraventricular arrhythmia | SNOMED | Arrythmia |
| 4064453 | 164893009 | EKG: ventricular arrhythmia | SNOMED | Arrythmia |
| 4064455 | 164896001 | EKG: ventricular fibrillation | SNOMED | Arrythmia |
| 4064869 | 164891006 | Electrocardiogram: paroxysmal atrial tachycardia | SNOMED | Arrythmia |
| 4226399 | 40593004 | Fibrillation | SNOMED | Arrythmia |
| 314379 | 270492004 | First degree atrioventricular block | SNOMED | Arrythmia |
| 4029303 | 13640000 | Fusion beats | SNOMED | Arrythmia |
| 320425 | 233916004 | Heart block | SNOMED | Arrythmia |
| 40481891 | 443478002 | Heart block caused by drug | SNOMED | Arrythmia |
| 43020494 | 284941000119107 | High degree second degree atrioventricular block | SNOMED | Arrythmia |
| 4120086 | 233901002 | His bundle tachycardia | SNOMED | Arrythmia |
| 46284985 | 959281000000105 | Holiday heart syndrome | SNOMED | Arrythmia |
| 4320474 | 69730002 | Idiojunctional tachycardia | SNOMED | Arrythmia |
| 37395937 | 715560009 | Idiopathic neonatal atrial flutter | SNOMED | Arrythmia |
| 37109917 | 723866006 | Idiopathic ventricular fibrillation not Brugada type | SNOMED | Arrythmia |
| 4171193 | 49260003 | Idioventricular rhythm | SNOMED | Arrythmia |
| 4124697 | 233894001 | Incessant atrial tachycardia | SNOMED | Arrythmia |
| 4298806 | 77221000 | Incomplete atrioventricular block with atrioventricular response | SNOMED | Arrythmia |
| 4088336 | 251120003 | Incomplete left bundle branch block | SNOMED | Arrythmia |
| 4088338 | 251124007 | Incomplete right bundle branch block | SNOMED | Arrythmia |
| 4088332 | 251114004 | Intermittent second degree atrioventricular block | SNOMED | Arrythmia |
| 4243143 | 38274001 | Interpolated ventricular premature complexes | SNOMED | Arrythmia |
| 4166844 | 4554005 | Intraventricular conduction defect | SNOMED | Arrythmia |
| 4221549 | 82838007 | Irregular tachycardia | SNOMED | Arrythmia |
| 4091899 | 251155001 | Junctional ectopic tachycardia | SNOMED | Arrythmia |
| 4166380 | 47830009 | Junctional escape beats | SNOMED | Arrythmia |
| 4218739 | 81681009 | Junctional premature beats | SNOMED | Arrythmia |
| 4088351 | 251164006 | Junctional premature complex | SNOMED | Arrythmia |
| 4295336 | 37760005 | Left anterior fascicular block | SNOMED | Arrythmia |
| 40482086 | 445118002 | Left anterior fascicular block on electrocardiogram | SNOMED | Arrythmia |
| 4171887 | 420002000 | Left atrial incisional tachycardia | SNOMED | Arrythmia |
| 316998 | 63467002 | Left bundle branch block | SNOMED | Arrythmia |
| 313209 | 4973001 | Left bundle branch hemiblock | SNOMED | Arrythmia |
| 4111543 | 195046004 | Left main stem bundle branch block | SNOMED | Arrythmia |
| 4268046 | 62026008 | Left posterior fascicular block | SNOMED | Arrythmia |
| 4153404 | 283645003 | Lev's syndrome | SNOMED | Arrythmia |
| 4119601 | 233910005 | Lone atrial fibrillation | SNOMED | Arrythmia |
| 314664 | 9651007 | Long QT syndrome | SNOMED | Arrythmia |
| 37396235 | 715971003 | Long QT syndrome caused by drug | SNOMED | Arrythmia |
| 45768480 | 706923002 | Longstanding persistent atrial fibrillation | SNOMED | Arrythmia |
| 42536724 | 735682000 | Macro re-entrant atrial tachycardia | SNOMED | Arrythmia |
| 46272503 | 710878005 | Mahaim fiber tachycardia | SNOMED | Arrythmia |
| 4088347 | 251152003 | Marked sinus arrhythmia | SNOMED | Arrythmia |
| 4088496 | 251125008 | Minor intraventricular conduction defect | SNOMED | Arrythmia |
| 313780 | 28189009 | Mobitz type II atrioventricular block | SNOMED | Arrythmia |
| 4205137 | 54016002 | Mobitz type I incomplete atrioventricular block | SNOMED | Arrythmia |
| 4064459 | 164905008 | Mobitz type I second degree atrioventricular block on electrocardiogram | SNOMED | Arrythmia |
| 4034164 | 2374000 | Monofascicular block | SNOMED | Arrythmia |
| 4176112 | 49982000 | Multifocal atrial tachycardia | SNOMED | Arrythmia |
| 4271464 | 63232000 | Multifocal premature beats | SNOMED | Arrythmia |
| 4023336 | 10626002 | Multifocal premature ventricular complexes | SNOMED | Arrythmia |
| 4089460 | 251171001 | Multiple atrial premature complexes | SNOMED | Arrythmia |
| 4088985 | 251176006 | Multiple premature ventricular complexes | SNOMED | Arrythmia |
| 4089463 | 251179004 | Multiple ventricular interpolated complexes | SNOMED | Arrythmia |
| 443522 | 413341007 | Neonatal bradycardia | SNOMED | Arrythmia |
| 4173170 | 276513001 | Neonatal dysrhythmia | SNOMED | Arrythmia |
| 443523 | 413342000 | Neonatal tachycardia | SNOMED | Arrythmia |
| 4217221 | 71792006 | Nodal rhythm disorder | SNOMED | Arrythmia |
| 42539038 | 735684004 | Non-cavotricuspid isthmus dependent atrial tachycardia | SNOMED | Arrythmia |
| 4191222 | 39260000 | Nonparoxysmal AV nodal tachycardia | SNOMED | Arrythmia |
| 4119602 | 233911009 | Non-rheumatic atrial fibrillation | SNOMED | Arrythmia |
| 44784220 | 698252002 | Non-specific intraventricular conduction delay | SNOMED | Arrythmia |
| 44782789 | 5761000119100 | Nonsustained paroxysmal supraventricular tachycardia | SNOMED | Arrythmia |
| 44782707 | 21421000119109 | Nonsustained paroxysmal ventricular tachycardia | SNOMED | Arrythmia |
| 4313053 | 423175003 | Normal sinus arrhythmia | SNOMED | Arrythmia |
| 4124700 | 233898003 | Orthodromic atrioventricular re-entrant tachycardia | SNOMED | Arrythmia |
| 4089464 | 251182009 | Paired ventricular premature complexes | SNOMED | Arrythmia |
| 4154290 | 282825002 | Paroxysmal atrial fibrillation | SNOMED | Arrythmia |
| 4146580 | 427665004 | Paroxysmal atrial flutter | SNOMED | Arrythmia |
| 4190306 | 39357005 | Paroxysmal atrial tachycardia with block | SNOMED | Arrythmia |
| 4108241 | 195070000 | Paroxysmal atrioventricular tachycardia | SNOMED | Arrythmia |
| 4111546 | 195071001 | Paroxysmal junctional tachycardia | SNOMED | Arrythmia |
| 4111698 | 195072008 | Paroxysmal nodal tachycardia | SNOMED | Arrythmia |
| 317893 | 67198005 | Paroxysmal supraventricular tachycardia | SNOMED | Arrythmia |
| 313792 | 12026006 | Paroxysmal tachycardia | SNOMED | Arrythmia |
| 437579 | 66657009 | Paroxysmal ventricular tachycardia | SNOMED | Arrythmia |
| 4111570 | 195039008 | Partial atrioventricular block | SNOMED | Arrythmia |
| 4232691 | 440028005 | Permanent atrial fibrillation | SNOMED | Arrythmia |
| 4124702 | 233904005 | Permanent junctional reciprocating tachycardia | SNOMED | Arrythmia |
| 4232697 | 440059007 | Persistent atrial fibrillation | SNOMED | Arrythmia |
| 4258998 | 44602002 | Persistent sinus bradycardia | SNOMED | Arrythmia |
| 43020495 | 284951000119109 | Postoperative atrioventricular block | SNOMED | Arrythmia |
| 4119605 | 233918003 | Postoperative complete heart block | SNOMED | Arrythmia |
| 4124701 | 233903004 | Postoperative His bundle tachycardia | SNOMED | Arrythmia |
| 4124704 | 233914001 | Postoperative sinus node dysfunction | SNOMED | Arrythmia |
| 42539346 | 762247006 | Preexcited atrial fibrillation | SNOMED | Arrythmia |
| 4109365 | 284470004 | Premature atrial contraction | SNOMED | Arrythmia |
| 316429 | 29717002 | Premature beats | SNOMED | Arrythmia |
| 44782643 | 698251009 | Progressive familial heart block, type II | SNOMED | Arrythmia |
| 4199501 | 314208002 | Rapid atrial fibrillation | SNOMED | Arrythmia |
| 4124696 | 233893007 | Re-entrant atrial tachycardia | SNOMED | Arrythmia |
| 4124698 | 233896004 | Re-entrant atrioventricular node tachycardia | SNOMED | Arrythmia |
| 4124699 | 233897008 | Re-entrant atrioventricular tachycardia | SNOMED | Arrythmia |
| 4111552 | 195105007 | Re-entry ventricular arrhythmia | SNOMED | Arrythmia |
| 4302802 | 418493005 | Right atrial incisional tachycardia | SNOMED | Arrythmia |
| 4249027 | 73459006 | Right branch block, incomplete anterior fascicular block AND incomplete posterior fascicular block | SNOMED | Arrythmia |
| 314059 | 59118001 | Right bundle branch block | SNOMED | Arrythmia |
| 4184950 | 43906007 | Right bundle branch block AND incomplete left bundle branch block | SNOMED | Arrythmia |
| 316432 | 30667004 | Right bundle branch block AND left anterior fascicular block | SNOMED | Arrythmia |
| 40482887 | 445263008 | Right bundle branch block and left anterior fascicular block on electrocardiogram | SNOMED | Arrythmia |
| 321590 | 46319007 | Right bundle branch block AND left posterior fascicular block | SNOMED | Arrythmia |
| 4244693 | 38566003 | Right bundle branch block, anterior fascicular block AND incomplete left bundle branch block | SNOMED | Arrythmia |
| 4217860 | 41863008 | Right bundle branch block, anterior fascicular block AND incomplete posterior fascicular block | SNOMED | Arrythmia |
| 4138545 | 32425009 | Right bundle branch block, anterior fascicular block AND posterior fascicular block | SNOMED | Arrythmia |
| 4280348 | 66568003 | Right bundle branch block, posterior fascicular block AND incomplete anterior fascicular block | SNOMED | Arrythmia |
| 4032785 | 14718009 | Right bundle branch block, posterior fascicular block AND incomplete left bundle branch block | SNOMED | Arrythmia |
| 4138973 | 32758004 | Right bundle branch block with left bundle branch block | SNOMED | Arrythmia |
| 4089461 | 251172008 | Run of atrial premature complexes | SNOMED | Arrythmia |
| 4091904 | 251177002 | Run of ventricular premature complexes | SNOMED | Arrythmia |
| 37312595 | 789039008 | Scar mediated macro re-entrant atrial tachycardia | SNOMED | Arrythmia |
| 318448 | 195042002 | Second degree atrioventricular block | SNOMED | Arrythmia |
| 4169261 | 49044005 | Severe sinus bradycardia | SNOMED | Arrythmia |
| 44784236 | 698272007 | Short QT syndrome | SNOMED | Arrythmia |
| 4261842 | 36083008 | Sick sinus syndrome | SNOMED | Arrythmia |
| 4028322 | 13395001 | Sinoatrial arrest with nodal/ventricular escape | SNOMED | Arrythmia |
| 4277903 | 65778007 | Sinoatrial block | SNOMED | Arrythmia |
| 4303256 | 419752005 | Sinoatrial nodal reentrant tachycardia | SNOMED | Arrythmia |
| 4120084 | 233891009 | Sinoatrial node tachycardia | SNOMED | Arrythmia |
| 4210313 | 5609005 | Sinus arrest | SNOMED | Arrythmia |
| 4088352 | 251092003 | Sinus arrest with atrial escape | SNOMED | Arrythmia |
| 4088210 | 251093008 | Sinus arrest with junctional escape | SNOMED | Arrythmia |
| 4091446 | 251094002 | Sinus arrest with ventricular escape | SNOMED | Arrythmia |
| 4171683 | 49710005 | Sinus bradycardia | SNOMED | Arrythmia |
| 317302 | 60423000 | Sinus node dysfunction | SNOMED | Arrythmia |
| 4088350 | 251161003 | Slow ventricular response | SNOMED | Arrythmia |
| 42536727 | 735686002 | Sudden arrhythmic death syndrome | SNOMED | Arrythmia |
| 4248028 | 72654001 | Supraventricular arrhythmia | SNOMED | Arrythmia |
| 4091902 | 251168009 | Supraventricular bigeminy | SNOMED | Arrythmia |
| 42538755 | 762534000 | Supraventricular bradyarrhythmia | SNOMED | Arrythmia |
| 441872 | 63593006 | Supraventricular premature beats | SNOMED | Arrythmia |
| 4275423 | 6456007 | Supraventricular tachycardia | SNOMED | Arrythmia |
| 4120085 | 233900001 | Supraventricular tachycardia with functional bundle branch block | SNOMED | Arrythmia |
| 4325850 | 429243003 | Sustained ventricular fibrillation | SNOMED | Arrythmia |
| 40480216 | 444605001 | Symptomatic sinus bradycardia | SNOMED | Arrythmia |
| 315643 | 6285003 | Tachyarrhythmia | SNOMED | Arrythmia |
| 4254116 | 74615001 | Tachycardia-bradycardia | SNOMED | Arrythmia |
| 4262389 | 46220003 | Tic-tac rhythm | SNOMED | Arrythmia |
| 321315 | 86014007 | Trifascicular block | SNOMED | Arrythmia |
| 40482938 | 445309007 | Trifascicular block on electrocardiogram | SNOMED | Arrythmia |
| 36714994 | 720448006 | Typical atrial flutter | SNOMED | Arrythmia |
| 4120087 | 233923003 | Unidirectional retrograde accessory pathway | SNOMED | Arrythmia |
| 4099778 | 27337007 | Unifocal premature ventricular complexes | SNOMED | Arrythmia |
| 4106715 | 29894000 | Vagal autonomic bradycardia | SNOMED | Arrythmia |
| 4185572 | 44103008 | Ventricular arrhythmia | SNOMED | Arrythmia |
| 4008580 | 11157007 | Ventricular bigeminy | SNOMED | Arrythmia |
| 4327066 | 75532003 | Ventricular escape beat | SNOMED | Arrythmia |
| 4088507 | 251186007 | Ventricular escape complex | SNOMED | Arrythmia |
| 4218242 | 81898007 | Ventricular escape rhythm | SNOMED | Arrythmia |
| 437894 | 71908006 | Ventricular fibrillation | SNOMED | Arrythmia |
| 4111700 | 195083004 | Ventricular fibrillation and flutter | SNOMED | Arrythmia |
| 433225 | 111288001 | Ventricular flutter | SNOMED | Arrythmia |
| 4244893 | 59272004 | Ventricular parasystole | SNOMED | Arrythmia |
| 4108828 | 195060002 | Ventricular pre-excitation | SNOMED | Arrythmia |
| 4089462 | 251175005 | Ventricular premature complex | SNOMED | Arrythmia |
| 4088506 | 251181002 | Ventricular quadrigeminy | SNOMED | Arrythmia |
| 40622721 | 6624005 | Ventricular tachyarrhythmia | SNOMED | Arrythmia |
| 4103295 | 25569003 | Ventricular tachycardia | SNOMED | Arrythmia |
| 4088505 | 251180001 | Ventricular trigeminy | SNOMED | Arrythmia |
| 4108832 | 195080001 | Atrial fibrillation and flutter | SNOMED | Arrythmia |
| 4089459 | 251166008 | AV nodal re-entry tachycardia | SNOMED | Arrythmia |
| 4092010 | 251178007 | Ventricular interpolated complexes | SNOMED | Arrythmia |
| 4066289 | 17338001 | Ventricular premature beats | SNOMED | Arrythmia |
| 4188347 | 46935006 | Stokes-Adams syndrome | SNOMED | Arrythmia |
| 762047 | 286411000119108 | Acute bilateral thrombosis of subclavian veins | SNOMED | Deep Vein Thrombosis (DVT) |
| 762148 | 293471000119106 | Acute deep vein thrombosis of bilateral iliac veins | SNOMED | Deep Vein Thrombosis (DVT) |
| 761444 | 15711361000119100 | Acute deep vein thrombosis of bilateral lower limbs following coronary artery bypass graft | SNOMED | Deep Vein Thrombosis (DVT) |
| 35616028 | 293491000119107 | Acute deep vein thrombosis of left iliac vein | SNOMED | Deep Vein Thrombosis (DVT) |
| 35615035 | 15711401000119100 | Acute deep vein thrombosis of left lower limb following procedure | SNOMED | Deep Vein Thrombosis (DVT) |
| 761416 | 15708441000119100 | Acute deep vein thrombosis of left upper limb following coronary artery bypass graft | SNOMED | Deep Vein Thrombosis (DVT) |
| 35615031 | 15708401000119100 | Acute deep vein thrombosis of left upper limb following procedure | SNOMED | Deep Vein Thrombosis (DVT) |
| 43531681 | 651000119108 | Acute deep vein thrombosis of lower limb | SNOMED | Deep Vein Thrombosis (DVT) |
| 35616027 | 293481000119109 | Acute deep vein thrombosis of right iliac vein | SNOMED | Deep Vein Thrombosis (DVT) |
| 35615034 | 15711241000119100 | Acute deep vein thrombosis of right lower limb following procedure | SNOMED | Deep Vein Thrombosis (DVT) |
| 761415 | 15708281000119100 | Acute deep vein thrombosis of right upper limb following coronary artery bypass graft | SNOMED | Deep Vein Thrombosis (DVT) |
| 35615030 | 15708201000119100 | Acute deep vein thrombosis of right upper limb following procedure | SNOMED | Deep Vein Thrombosis (DVT) |
| 44782746 | 132281000119108 | Acute deep venous thrombosis | SNOMED | Deep Vein Thrombosis (DVT) |
| 44782751 | 134961000119104 | Acute deep venous thrombosis of axillary vein | SNOMED | Deep Vein Thrombosis (DVT) |
| 762008 | 285321000119103 | Acute deep venous thrombosis of bilateral axillary veins | SNOMED | Deep Vein Thrombosis (DVT) |
| 760875 | 12237551000119100 | Acute deep venous thrombosis of bilateral calves | SNOMED | Deep Vein Thrombosis (DVT) |
| 765155 | 285441000119102 | Acute deep venous thrombosis of bilateral ileofemoral veins | SNOMED | Deep Vein Thrombosis (DVT) |
| 762017 | 285501000119103 | Acute deep venous thrombosis of bilateral internal jugular veins | SNOMED | Deep Vein Thrombosis (DVT) |
| 762417 | 350291000119100 | Acute deep venous thrombosis of bilateral legs | SNOMED | Deep Vein Thrombosis (DVT) |
| 762020 | 285561000119102 | Acute deep venous thrombosis of bilateral popliteal veins | SNOMED | Deep Vein Thrombosis (DVT) |
| 765546 | 285621000119106 | Acute deep venous thrombosis of bilateral tibial veins | SNOMED | Deep Vein Thrombosis (DVT) |
| 762004 | 285261000119104 | Acute deep venous thrombosis of both upper extremities | SNOMED | Deep Vein Thrombosis (DVT) |
| 44782742 | 132241000119103 | Acute deep venous thrombosis of calf | SNOMED | Deep Vein Thrombosis (DVT) |
| 44782747 | 132291000119106 | Acute deep venous thrombosis of femoral vein | SNOMED | Deep Vein Thrombosis (DVT) |
| 762015 | 285451000119100 | Acute deep venous thrombosis of ileofemoral vein of left leg | SNOMED | Deep Vein Thrombosis (DVT) |
| 765541 | 285461000119103 | Acute deep venous thrombosis of ileofemoral vein of right lower extremity | SNOMED | Deep Vein Thrombosis (DVT) |
| 44782748 | 132301000119107 | Acute deep venous thrombosis of iliofemoral vein | SNOMED | Deep Vein Thrombosis (DVT) |
| 44782752 | 135001000119100 | Acute deep venous thrombosis of internal jugular vein | SNOMED | Deep Vein Thrombosis (DVT) |
| 762009 | 285331000119100 | Acute deep venous thrombosis of left axillary vein | SNOMED | Deep Vein Thrombosis (DVT) |
| 760876 | 12237631000119100 | Acute deep venous thrombosis of left calf | SNOMED | Deep Vein Thrombosis (DVT) |
| 765540 | 285391000119101 | Acute deep venous thrombosis of left femoral vein | SNOMED | Deep Vein Thrombosis (DVT) |
| 765922 | 285511000119100 | Acute deep venous thrombosis of left internal jugular vein | SNOMED | Deep Vein Thrombosis (DVT) |
| 762418 | 350301000119104 | Acute deep venous thrombosis of left lower extremity | SNOMED | Deep Vein Thrombosis (DVT) |
| 765537 | 285271000119105 | Acute deep venous thrombosis of left upper extremity | SNOMED | Deep Vein Thrombosis (DVT) |
| 44782767 | 136781000119101 | Acute deep venous thrombosis of lower extremity as complication of procedure | SNOMED | Deep Vein Thrombosis (DVT) |
| 46270071 | 132111000119107 | Acute deep venous thrombosis of lower limb due to and following coronary artery bypass grafting | SNOMED | Deep Vein Thrombosis (DVT) |
| 762022 | 285581000119106 | Acute deep venous thrombosis of politeal vein of right leg | SNOMED | Deep Vein Thrombosis (DVT) |
| 44782743 | 132251000119101 | Acute deep venous thrombosis of popliteal vein | SNOMED | Deep Vein Thrombosis (DVT) |
| 762021 | 285571000119108 | Acute deep venous thrombosis of popliteal vein of left leg | SNOMED | Deep Vein Thrombosis (DVT) |
| 762010 | 285341000119109 | Acute deep venous thrombosis of right axillary vein | SNOMED | Deep Vein Thrombosis (DVT) |
| 760877 | 12237711000119100 | Acute deep venous thrombosis of right calf | SNOMED | Deep Vein Thrombosis (DVT) |
| 762013 | 285401000119104 | Acute deep venous thrombosis of right femoral vein | SNOMED | Deep Vein Thrombosis (DVT) |
| 762018 | 285521000119107 | Acute deep venous thrombosis of right internal jugular vein | SNOMED | Deep Vein Thrombosis (DVT) |
| 762419 | 350311000119101 | Acute deep venous thrombosis of right lower extremity | SNOMED | Deep Vein Thrombosis (DVT) |
| 762005 | 285281000119108 | Acute deep venous thrombosis of right upper extremity | SNOMED | Deep Vein Thrombosis (DVT) |
| 44782745 | 132271000119105 | Acute deep venous thrombosis of thigh | SNOMED | Deep Vein Thrombosis (DVT) |
| 44782744 | 132261000119104 | Acute deep venous thrombosis of tibial vein | SNOMED | Deep Vein Thrombosis (DVT) |
| 762026 | 285631000119109 | Acute deep venous thrombosis of tibial vein of left leg | SNOMED | Deep Vein Thrombosis (DVT) |
| 765156 | 285641000119100 | Acute deep venous thrombosis of tibial vein of right leg | SNOMED | Deep Vein Thrombosis (DVT) |
| 44782421 | 132321000119103 | Acute deep venous thrombosis of upper extremity | SNOMED | Deep Vein Thrombosis (DVT) |
| 764016 | 449691000124103 | Acute deep venous thrombosis of upper extremity after coronary artery bypass graft | SNOMED | Deep Vein Thrombosis (DVT) |
| 44782766 | 136771000119104 | Acute deep venous thrombosis of upper extremity as complication of procedure | SNOMED | Deep Vein Thrombosis (DVT) |
| 762048 | 286421000119101 | Acute thrombosis of left subclavian vein | SNOMED | Deep Vein Thrombosis (DVT) |
| 45757410 | 133421000119101 | Acute thrombosis of mesenteric vein | SNOMED | Deep Vein Thrombosis (DVT) |
| 762049 | 286431000119103 | Acute thrombosis of right subclavian vein | SNOMED | Deep Vein Thrombosis (DVT) |
| 36712892 | 143561000119108 | Acute thrombosis of splenic vein | SNOMED | Deep Vein Thrombosis (DVT) |
| 44782762 | 132611000119104 | Acute thrombosis of subclavian vein | SNOMED | Deep Vein Thrombosis (DVT) |
| 435887 | 49956009 | Antepartum deep vein thrombosis | SNOMED | Deep Vein Thrombosis (DVT) |
| 4179911 | 297156001 | Axillary vein thrombosis | SNOMED | Deep Vein Thrombosis (DVT) |
| 37109253 | 285381000119104 | Bilateral acute deep vein thrombosis of femoral veins | SNOMED | Deep Vein Thrombosis (DVT) |
| 40478951 | 444325005 | Bilateral deep vein thrombosis of lower extremities | SNOMED | Deep Vein Thrombosis (DVT) |
| 4042396 | 16750002 | Deep thrombophlebitis | SNOMED | Deep Vein Thrombosis (DVT) |
| 4046884 | 134399007 | Deep vein thrombosis of leg related to air travel | SNOMED | Deep Vein Thrombosis (DVT) |
| 3655221 | 860699005 | Deep vein thrombosis of lower extremity due to intravenous drug use | SNOMED | Deep Vein Thrombosis (DVT) |
| 4133004 | 128053003 | Deep venous thrombosis | SNOMED | Deep Vein Thrombosis (DVT) |
| 4181315 | 428781001 | Deep venous thrombosis associated with coronary artery bypass graft | SNOMED | Deep Vein Thrombosis (DVT) |
| 438820 | 56272000 | Deep venous thrombosis in puerperium | SNOMED | Deep Vein Thrombosis (DVT) |
| 45773536 | 703277001 | Deep venous thrombosis of femoropopliteal vein | SNOMED | Deep Vein Thrombosis (DVT) |
| 763942 | 448841000124100 | Deep venous thrombosis of left lower extremity | SNOMED | Deep Vein Thrombosis (DVT) |
| 761980 | 25820001000004100 | Deep venous thrombosis of left upper extremity | SNOMED | Deep Vein Thrombosis (DVT) |
| 443537 | 404223003 | Deep venous thrombosis of lower extremity | SNOMED | Deep Vein Thrombosis (DVT) |
| 4133975 | 128055005 | Deep venous thrombosis of pelvic vein | SNOMED | Deep Vein Thrombosis (DVT) |
| 40480555 | 443210003 | Deep venous thrombosis of peroneal vein | SNOMED | Deep Vein Thrombosis (DVT) |
| 4322565 | 427775006 | Deep venous thrombosis of profunda femoris vein | SNOMED | Deep Vein Thrombosis (DVT) |
| 763941 | 448831000124105 | Deep venous thrombosis of right lower extremity | SNOMED | Deep Vein Thrombosis (DVT) |
| 761928 | 20850001000004100 | Deep venous thrombosis of right upper extremity | SNOMED | Deep Vein Thrombosis (DVT) |
| 4207899 | 438785004 | Deep venous thrombosis of tibial vein | SNOMED | Deep Vein Thrombosis (DVT) |
| 4028057 | 128054009 | Deep venous thrombosis of upper extremity | SNOMED | Deep Vein Thrombosis (DVT) |
| 193512 | 195438008 | Embolism and thrombosis of the renal vein | SNOMED | Deep Vein Thrombosis (DVT) |
| 435565 | 195437003 | Embolism and thrombosis of the vena cava | SNOMED | Deep Vein Thrombosis (DVT) |
| 40481089 | 444816006 | Embolism from thrombosis of vein of lower extremity | SNOMED | Deep Vein Thrombosis (DVT) |
| 4119760 | 234044007 | Iliofemoral deep vein thrombosis | SNOMED | Deep Vein Thrombosis (DVT) |
| 4124856 | 234041004 | Inferior mesenteric vein thrombosis | SNOMED | Deep Vein Thrombosis (DVT) |
| 4096099 | 25114006 | Phlebitis of deep veins of lower extremity | SNOMED | Deep Vein Thrombosis (DVT) |
| 4111861 | 195405004 | Phlebitis of popliteal vein | SNOMED | Deep Vein Thrombosis (DVT) |
| 440738 | 195404000 | Phlebitis of the femoral vein | SNOMED | Deep Vein Thrombosis (DVT) |
| 4281689 | 66923004 | Phlegmasia alba dolens | SNOMED | Deep Vein Thrombosis (DVT) |
| 4284538 | 66877004 | Phlegmasia cerulea dolens | SNOMED | Deep Vein Thrombosis (DVT) |
| 199837 | 17920008 | Portal vein thrombosis | SNOMED | Deep Vein Thrombosis (DVT) |
| 4309333 | 213220000 | Postoperative deep vein thrombosis | SNOMED | Deep Vein Thrombosis (DVT) |
| 46285905 | 978441000000108 | Provoked deep vein thrombosis | SNOMED | Deep Vein Thrombosis (DVT) |
| 46271900 | 710167004 | Recurrent deep vein thrombosis | SNOMED | Deep Vein Thrombosis (DVT) |
| 4033521 | 14534009 | Splenic vein thrombosis | SNOMED | Deep Vein Thrombosis (DVT) |
| 4055089 | 197001004 | Superior mesenteric vein thrombosis | SNOMED | Deep Vein Thrombosis (DVT) |
| 4230403 | 438646004 | Thrombophlebitis of axillary vein | SNOMED | Deep Vein Thrombosis (DVT) |
| 4069561 | 1748006 | Thrombophlebitis of deep femoral vein | SNOMED | Deep Vein Thrombosis (DVT) |
| 761831 | 16014391000119100 | Thrombophlebitis of deep vein of bilateral lower limbs | SNOMED | Deep Vein Thrombosis (DVT) |
| 761830 | 16014351000119100 | Thrombophlebitis of deep vein of left lower limb | SNOMED | Deep Vein Thrombosis (DVT) |
| 761808 | 16006271000119100 | Thrombophlebitis of deep vein of left upper limb | SNOMED | Deep Vein Thrombosis (DVT) |
| 761832 | 16014431000119100 | Thrombophlebitis of deep vein of right lower limb | SNOMED | Deep Vein Thrombosis (DVT) |
| 761809 | 16006311000119100 | Thrombophlebitis of deep vein of right upper limb | SNOMED | Deep Vein Thrombosis (DVT) |
| 4221821 | 40198004 | Thrombophlebitis of deep veins of lower extremity | SNOMED | Deep Vein Thrombosis (DVT) |
| 440750 | 95452006 | Thrombophlebitis of deep veins of upper extremities | SNOMED | Deep Vein Thrombosis (DVT) |
| 4176614 | 42861008 | Thrombophlebitis of iliac vein | SNOMED | Deep Vein Thrombosis (DVT) |
| 761821 | 16012151000119100 | Thrombophlebitis of left deep femoral vein | SNOMED | Deep Vein Thrombosis (DVT) |
| 761819 | 16012071000119100 | Thrombophlebitis of left femoral vein | SNOMED | Deep Vein Thrombosis (DVT) |
| 761820 | 16012111000119100 | Thrombophlebitis of right deep femoral vein | SNOMED | Deep Vein Thrombosis (DVT) |
| 761818 | 16011991000119100 | Thrombophlebitis of right femoral vein | SNOMED | Deep Vein Thrombosis (DVT) |
| 4110339 | 195412008 | Thrombophlebitis of the anterior tibial vein | SNOMED | Deep Vein Thrombosis (DVT) |
| 4111868 | 195425000 | Thrombophlebitis of the common iliac vein | SNOMED | Deep Vein Thrombosis (DVT) |
| 4110343 | 195427008 | Thrombophlebitis of the external iliac vein | SNOMED | Deep Vein Thrombosis (DVT) |
| 439314 | 195410000 | Thrombophlebitis of the femoral vein | SNOMED | Deep Vein Thrombosis (DVT) |
| 4109877 | 195426004 | Thrombophlebitis of the internal iliac vein | SNOMED | Deep Vein Thrombosis (DVT) |
| 4112171 | 195411001 | Thrombophlebitis of the popliteal vein | SNOMED | Deep Vein Thrombosis (DVT) |
| 4112172 | 195414009 | Thrombophlebitis of the posterior tibial vein | SNOMED | Deep Vein Thrombosis (DVT) |
| 4250765 | 7387004 | Thrombophlebitis of tibial vein | SNOMED | Deep Vein Thrombosis (DVT) |
| 42538533 | 762256003 | Thrombosis of iliac vein | SNOMED | Deep Vein Thrombosis (DVT) |
| 44811347 | 864191000000104 | Thrombosis of internal jugular vein | SNOMED | Deep Vein Thrombosis (DVT) |
| 765049 | 16730001000004100 | Thrombosis of left peroneal vein | SNOMED | Deep Vein Thrombosis (DVT) |
| 4317289 | 95446005 | Thrombosis of mesenteric vein | SNOMED | Deep Vein Thrombosis (DVT) |
| 4203836 | 438647008 | Thrombosis of subclavian vein | SNOMED | Deep Vein Thrombosis (DVT) |
| 4175649 | 427776007 | Thrombosis of the popliteal vein | SNOMED | Deep Vein Thrombosis (DVT) |
| 4149782 | 309735004 | Thrombosis of vein of lower limb | SNOMED | Deep Vein Thrombosis (DVT) |
| 4153353 | 371051005 | Traumatic thrombosis of axillary vein | SNOMED | Deep Vein Thrombosis (DVT) |
| 46285904 | 978421000000101 | Unprovoked deep vein thrombosis | SNOMED | Deep Vein Thrombosis (DVT) |
| 77310 | 266267005 | Deep vein phlebitis and thrombophlebitis of the leg | SNOMED | Deep Vein Thrombosis (DVT) |
| 4189004 | 413956008 | Deep vein thrombosis of leg related to intravenous drug use | SNOMED | Deep Vein Thrombosis (DVT) |
| 4172545 | 277303004 | Angiogram-negative subarachnoid hemorrhage | SNOMED | Hemorrhagic stroke |
| 4112018 | 195165005 | Basal ganglia hemorrhage | SNOMED | Hemorrhagic stroke |
| 4319328 | 95454007 | Brain stem hemorrhage | SNOMED | Hemorrhagic stroke |
| 4201094 | 301765007 | Cerebellar hematoma | SNOMED | Hemorrhagic stroke |
| 4326561 | 75038005 | Cerebellar hemorrhage | SNOMED | Hemorrhagic stroke |
| 4218781 | 73020009 | Cerebral hemisphere hemorrhage | SNOMED | Hemorrhagic stroke |
| 376713 | 274100004 | Cerebral hemorrhage | SNOMED | Hemorrhagic stroke |
| 4173481 | 42429001 | Cerebromeningeal hemorrhage | SNOMED | Hemorrhagic stroke |
| 45766120 | 703217005 | Convexal subarachnoid hemorrhage | SNOMED | Hemorrhagic stroke |
| 4176892 | 49422009 | Cortical hemorrhage | SNOMED | Hemorrhagic stroke |
| 45766068 | 703156006 | Deep hemispheric cerebral hemorrhage | SNOMED | Hemorrhagic stroke |
| 4112019 | 195167002 | External capsule hemorrhage | SNOMED | Hemorrhagic stroke |
| 35609033 | 1078001000000100 | Haemorrhagic stroke | SNOMED | Hemorrhagic stroke |
| 4199890 | 301764006 | Hematoma of brain | SNOMED | Hemorrhagic stroke |
| 42873045 | 451038000 | Hemorrhage in caudate nucleus | SNOMED | Hemorrhagic stroke |
| 42873044 | 451037005 | Hemorrhage in globus pallidus | SNOMED | Hemorrhagic stroke |
| 42873046 | 451039008 | Hemorrhage in putamen | SNOMED | Hemorrhagic stroke |
| 37116267 | 732923001 | Hemorrhage of medulla oblongata | SNOMED | Hemorrhagic stroke |
| 4178726 | 52201006 | Internal capsule hemorrhage | SNOMED | Hemorrhagic stroke |
| 4080892 | 276722003 | Intracerebellar and posterior fossa hemorrhage | SNOMED | Hemorrhagic stroke |
| 4110185 | 195168007 | Intracerebral hemorrhage, intraventricular | SNOMED | Hemorrhagic stroke |
| 4110186 | 195169004 | Intracerebral hemorrhage, multiple localized | SNOMED | Hemorrhagic stroke |
| 42872434 | 450425005 | Intracranial hematoma | SNOMED | Hemorrhagic stroke |
| 439847 | 1386000 | Intracranial hemorrhage | SNOMED | Hemorrhagic stroke |
| 4328027 | 431266005 | Intraparenchymal hematoma of brain | SNOMED | Hemorrhagic stroke |
| 40492969 | 449020009 | Intraparenchymal hemorrhage of brain | SNOMED | Hemorrhagic stroke |
| 4299377 | 7713009 | Intrapontine hemorrhage | SNOMED | Hemorrhagic stroke |
| 4045744 | 230710000 | Lobar cerebral hemorrhage | SNOMED | Hemorrhagic stroke |
| 4045743 | 230709005 | Massive supratentorial cerebral hemorrhage | SNOMED | Hemorrhagic stroke |
| 45766830 | 704079000 | Non-aneurysmal perimesencephalic subarachnoid hemorrhage | SNOMED | Hemorrhagic stroke |
| 45766118 | 703215002 | Non-aneurysmal subarachnoid intracranial hemorrhage | SNOMED | Hemorrhagic stroke |
| 762096 | 291471000119107 | Non-traumatic hemorrhage of subarachnoid space from right middle cerebral artery | SNOMED | Hemorrhagic stroke |
| 4144154 | 425957003 | Non-traumatic intracerebral ventricular hemorrhage | SNOMED | Hemorrhagic stroke |
| 44782730 | 143521000119103 | Nontraumatic intraparenchymal cerebral hemorrhage | SNOMED | Hemorrhagic stroke |
| 46270111 | 141091000119105 | Nontraumatic subarachnoid hemorrhage with brain compression | SNOMED | Hemorrhagic stroke |
| 45766119 | 703216001 | Perimesencephalic subarachnoid hemorrhage | SNOMED | Hemorrhagic stroke |
| 4298750 | 384993003 | Periventricular hemorrhagic venous infarct | SNOMED | Hemorrhagic stroke |
| 4129535 | 237702003 | Pituitary hemorrhage | SNOMED | Hemorrhagic stroke |
| 762351 | 330111000119103 | Ruptured acquired aneurysm of cerebral artery | SNOMED | Hemorrhagic stroke |
| 764707 | 5471000124102 | Ruptured aneurysm of intracranial artery | SNOMED | Hemorrhagic stroke |
| 37109909 | 723857007 | Silent micro-hemorrhage of brain | SNOMED | Hemorrhagic stroke |
| 43530674 | 142851000119103 | Spontaneous cerebellar hemorrhage | SNOMED | Hemorrhagic stroke |
| 43530727 | 291571000119106 | Spontaneous cerebral hemorrhage | SNOMED | Hemorrhagic stroke |
| 46273491 | 140881000119109 | Spontaneous cerebral hemorrhage with compression of brain | SNOMED | Hemorrhagic stroke |
| 42539269 | 291541000119104 | Spontaneous hemorrhage of brain stem | SNOMED | Hemorrhagic stroke |
| 42535425 | 291531000119108 | Spontaneous hemorrhage of cerebral hemisphere | SNOMED | Hemorrhagic stroke |
| 42535424 | 291521000119105 | Spontaneous hemorrhage of cortical intracerebral hemisphere | SNOMED | Hemorrhagic stroke |
| 42535423 | 291511000119103 | Spontaneous hemorrhage of deep cerebral hemisphere | SNOMED | Hemorrhagic stroke |
| 37309659 | 16709811000119100 | Spontaneous hemorrhage of subarachnoid space from anterior communicating artery | SNOMED | Hemorrhagic stroke |
| 42535420 | 291351000119109 | Spontaneous hemorrhage of subarachnoid space from basilar artery | SNOMED | Hemorrhagic stroke |
| 42535421 | 291371000119100 | Spontaneous hemorrhage of subarachnoid space from intracranial artery | SNOMED | Hemorrhagic stroke |
| 762095 | 291401000119102 | Spontaneous hemorrhage of subarachnoid space from left middle cerebral artery | SNOMED | Hemorrhagic stroke |
| 42535422 | 291411000119104 | Spontaneous hemorrhage of subarachnoid space from left posterior communicating artery | SNOMED | Hemorrhagic stroke |
| 42539183 | 291481000119105 | Spontaneous hemorrhage of subarachnoid space from right posterior communicating artery | SNOMED | Hemorrhagic stroke |
| 42538062 | 738779002 | Spontaneous intracranial hemorrhage | SNOMED | Hemorrhagic stroke |
| 4148906 | 270907008 | Spontaneous subarachnoid hemorrhage | SNOMED | Hemorrhagic stroke |
| 432923 | 21454007 | Subarachnoid hemorrhage | SNOMED | Hemorrhagic stroke |
| 4046365 | 230719004 | Subarachnoid hemorrhage due to ruptured aneurysm | SNOMED | Hemorrhagic stroke |
| 4046364 | 230718007 | Subarachnoid hemorrhage due to ruptured arteriovenous malformation | SNOMED | Hemorrhagic stroke |
| 4077828 | 276278003 | Subarachnoid hemorrhage from anterior cerebral artery aneurysm | SNOMED | Hemorrhagic stroke |
| 4077958 | 276282001 | Subarachnoid hemorrhage from anterior communicating artery aneurysm | SNOMED | Hemorrhagic stroke |
| 4077201 | 276284000 | Subarachnoid hemorrhage from basilar artery aneurysm | SNOMED | Hemorrhagic stroke |
| 4078448 | 276286003 | Subarachnoid hemorrhage from carotid artery aneurysm | SNOMED | Hemorrhagic stroke |
| 4108952 | 195155004 | Subarachnoid hemorrhage from carotid siphon and bifurcation | SNOMED | Hemorrhagic stroke |
| 4078446 | 276280009 | Subarachnoid hemorrhage from middle cerebral artery aneurysm | SNOMED | Hemorrhagic stroke |
| 4077827 | 276277008 | Subarachnoid hemorrhage from multiple aneurysms | SNOMED | Hemorrhagic stroke |
| 4077200 | 276281008 | Subarachnoid hemorrhage from posterior cerebral artery aneurysm | SNOMED | Hemorrhagic stroke |
| 4077959 | 276283006 | Subarachnoid hemorrhage from posterior communicating artery aneurysm | SNOMED | Hemorrhagic stroke |
| 4078447 | 276285004 | Subarachnoid hemorrhage from posterior inferior cerebellar artery aneurysm | SNOMED | Hemorrhagic stroke |
| 4111708 | 195160000 | Subarachnoid hemorrhage from vertebral artery | SNOMED | Hemorrhagic stroke |
| 45773167 | 703174002 | Subarachnoid hemorrhage from vertebral artery aneurysm | SNOMED | Hemorrhagic stroke |
| 4049659 | 20908003 | Subcortical hemorrhage | SNOMED | Hemorrhagic stroke |
| 42873042 | 451035002 | Subpial intracranial hemorrhage | SNOMED | Hemorrhagic stroke |
| 4045745 | 230711001 | Thalamic hemorrhage | SNOMED | Hemorrhagic stroke |
| 443752 | 23276006 | Ventricular hemorrhage | SNOMED | Hemorrhagic stroke |
| 44782718 | 153931000119109 | Acute combined systolic and diastolic heart failure | SNOMED | Heart failure |
| 4023479 | 10633002 | Acute congestive heart failure | SNOMED | Heart failure |
| 312927 | 49584005 | Acute cor pulmonale | SNOMED | Heart failure |
| 40481042 | 443343001 | Acute diastolic heart failure | SNOMED | Heart failure |
| 44782655 | 698296002 | Acute exacerbation of chronic congestive heart failure | SNOMED | Heart failure |
| 442310 | 56675007 | Acute heart failure | SNOMED | Heart failure |
| 764877 | 7421000175106 | Acute heart failure co-occurrent with normal ejection fraction | SNOMED | Heart failure |
| 4327205 | 74960003 | Acute left-sided congestive heart failure | SNOMED | Heart failure |
| 4267800 | 364006 | Acute left-sided heart failure | SNOMED | Heart failure |
| 4108245 | 195114002 | Acute left ventricular failure | SNOMED | Heart failure |
| 44782733 | 153951000119103 | Acute on chronic combined systolic and diastolic heart failure | SNOMED | Heart failure |
| 40481043 | 443344007 | Acute on chronic diastolic heart failure | SNOMED | Heart failure |
| 764874 | 7401000175100 | Acute on chronic heart failure co-occurrent with normal ejection fraction | SNOMED | Heart failure |
| 37309625 | 16838951000119100 | Acute on chronic right-sided congestive heart failure | SNOMED | Heart failure |
| 40480602 | 443253003 | Acute on chronic systolic heart failure | SNOMED | Heart failure |
| 4215446 | 80479009 | Acute right-sided congestive heart failure | SNOMED | Heart failure |
| 4233424 | 359617009 | Acute right-sided heart failure | SNOMED | Heart failure |
| 40480603 | 443254009 | Acute systolic heart failure | SNOMED | Heart failure |
| 439698 | 194767001 | Benign hypertensive heart disease with congestive cardiac failure | SNOMED | Heart failure |
| 4242669 | 92506005 | Biventricular congestive heart failure | SNOMED | Heart failure |
| 4205558 | 55565007 | Cardiac failure after obstetrical surgery AND/OR other procedure including delivery | SNOMED | Heart failure |
| 4146456 | 308118002 | Cardiac failure therapy | SNOMED | Heart failure |
| 4233224 | 89819002 | Cardiac insufficiency during AND/OR resulting from a procedure | SNOMED | Heart failure |
| 4264636 | 60856006 | Cardiac insufficiency following cardiac surgery | SNOMED | Heart failure |
| 321319 | 85898001 | Cardiomyopathy | SNOMED | Heart failure |
| 40482857 | 445236007 | Cardiorenal syndrome | SNOMED | Heart failure |
| 4259490 | 410431009 | Cardiorespiratory failure | SNOMED | Heart failure |
| 44782719 | 153941000119100 | Chronic combined systolic and diastolic heart failure | SNOMED | Heart failure |
| 4229440 | 88805009 | Chronic congestive heart failure | SNOMED | Heart failure |
| 4195892 | 79955004 | Chronic cor pulmonale | SNOMED | Heart failure |
| 40479576 | 441530006 | Chronic diastolic heart failure | SNOMED | Heart failure |
| 444031 | 48447003 | Chronic heart failure | SNOMED | Heart failure |
| 764876 | 7411000175102 | Chronic heart failure co-occurrent with normal ejection fraction | SNOMED | Heart failure |
| 4206009 | 5375005 | Chronic left-sided congestive heart failure | SNOMED | Heart failure |
| 4009047 | 111283005 | Chronic left-sided heart failure | SNOMED | Heart failure |
| 4284562 | 66989003 | Chronic right-sided congestive heart failure | SNOMED | Heart failure |
| 4014159 | 10335000 | Chronic right-sided heart failure | SNOMED | Heart failure |
| 40479192 | 441481004 | Chronic systolic heart failure | SNOMED | Heart failure |
| 4108244 | 195112003 | Compensated cardiac failure | SNOMED | Heart failure |
| 4071869 | 206586007 | Congenital cardiac failure | SNOMED | Heart failure |
| 319835 | 42343007 | Congestive heart failure | SNOMED | Heart failure |
| 44784345 | 72481000119103 | Congestive heart failure as early postoperative complication | SNOMED | Heart failure |
| 762002 | 285211000119102 | Congestive heart failure as post-operative complication of cardiac surgery | SNOMED | Heart failure |
| 762003 | 285221000119109 | Congestive heart failure as post-operative complication of non-cardiac surgery | SNOMED | Heart failure |
| 44782428 | 101281000119107 | Congestive heart failure due to cardiomyopathy | SNOMED | Heart failure |
| 4139864 | 426263006 | Congestive heart failure due to left ventricular systolic dysfunction | SNOMED | Heart failure |
| 4142561 | 426611007 | Congestive heart failure due to valvular disease | SNOMED | Heart failure |
| 36712928 | 15629591000119100 | Congestive heart failure stage B due to ischemic cardiomyopathy | SNOMED | Heart failure |
| 43021826 | 67441000119101 | Congestive heart failure stage C | SNOMED | Heart failure |
| 36712927 | 15629541000119100 | Congestive heart failure stage C due to ischemic cardiomyopathy | SNOMED | Heart failure |
| 43021825 | 67431000119105 | Congestive heart failure stage D | SNOMED | Heart failure |
| 44782713 | 23341000119109 | Congestive heart failure with right heart failure | SNOMED | Heart failure |
| 315295 | 82523003 | Congestive rheumatic heart failure | SNOMED | Heart failure |
| 4307356 | 83291003 | Cor pulmonale | SNOMED | Heart failure |
| 4111554 | 195111005 | Decompensated cardiac failure | SNOMED | Heart failure |
| 4311437 | 424404003 | Decompensated chronic heart failure | SNOMED | Heart failure |
| 443587 | 418304008 | Diastolic heart failure | SNOMED | Heart failure |
| 43021842 | 120891000119109 | Diastolic heart failure stage C | SNOMED | Heart failure |
| 43021841 | 120881000119106 | Diastolic heart failure stage D | SNOMED | Heart failure |
| 4163710 | 399020009 | Dilated cardiomyopathy | SNOMED | Heart failure |
| 44805683 | 765661000000102 | Discharge from heart failure nurse service | SNOMED | Heart failure |
| 4215511 | 416683003 | Emergency hospital admission for heart failure | SNOMED | Heart failure |
| 43022068 | 96311000119109 | Exacerbation of congestive heart failure | SNOMED | Heart failure |
| 36204172 | 86490-0 | Felt discouraged or down in the dumps because of heart failure over the past 2 weeks [KCCQ] | LOINC | Heart failure |
| 4110961 | 194849004 | Generalized ischemic myocardial dysfunction | SNOMED | Heart failure |
| 44813162 | 781051000000108 | Has heart failure management plan | SNOMED | Heart failure |
| 316139 | 84114007 | Heart failure | SNOMED | Heart failure |
| 44790972 | 247361000000100 | Heart failure 6 month review | SNOMED | Heart failure |
| 4193010 | 390885007 | Heart failure annual review | SNOMED | Heart failure |
| 4124705 | 233924009 | Heart failure as a complication of care | SNOMED | Heart failure |
| 44808957 | 851521000000102 | Heart failure clinical pathway | SNOMED | Heart failure |
| 4215689 | 395105005 | Heart failure confirmed | SNOMED | Heart failure |
| 43020657 | 471880001 | Heart failure due to end stage congenital heart disease | SNOMED | Heart failure |
| 44808862 | 851071000000108 | Heart failure initial assessment | SNOMED | Heart failure |
| 44789102 | 202231000000106 | Heart failure review completed | SNOMED | Heart failure |
| 44806371 | 812001000000102 | Heart failure self-management plan agreed | SNOMED | Heart failure |
| 44806125 | 810971000000105 | Heart failure self-management plan review | SNOMED | Heart failure |
| 37311948 | 788950000 | Heart failure with mid range ejection fraction | SNOMED | Heart failure |
| 40486933 | 446221000 | Heart failure with normal ejection fraction | SNOMED | Heart failure |
| 45766164 | 703272007 | Heart failure with reduced ejection fraction | SNOMED | Heart failure |
| 45766167 | 703275009 | Heart failure with reduced ejection fraction due to cardiomyopathy | SNOMED | Heart failure |
| 45766165 | 703273002 | Heart failure with reduced ejection fraction due to coronary artery disease | SNOMED | Heart failure |
| 45773075 | 703276005 | Heart failure with reduced ejection fraction due to heart valve disease | SNOMED | Heart failure |
| 45766166 | 703274008 | Heart failure with reduced ejection fraction due to myocarditis | SNOMED | Heart failure |
| 4004279 | 10091002 | High output heart failure | SNOMED | Heart failure |
| 36305423 | 88653-1 | History of Acute heart failure within 24 hours prior to procedure | LOINC | Heart failure |
| 36204171 | 86489-2 | How would you feel if you had to spend the rest of your life with your heart failure the way it is right now [KCCQ] | LOINC | Heart failure |
| 36204159 | 86477-7 | Hurrying or jogging (as if to catch a bus) was limited by heart failure over the past 2 weeks [KCCQ] | LOINC | Heart failure |
| 44784621 | 8501000119104 | Hypertensive heart and chronic kidney disease | SNOMED | Heart failure |
| 44782728 | 15781000119107 | Hypertensive heart AND chronic kidney disease with congestive heart failure | SNOMED | Heart failure |
| 439694 | 194781004 | Hypertensive heart and renal disease with both (congestive) heart failure and renal failure | SNOMED | Heart failure |
| 439696 | 194779001 | Hypertensive heart and renal disease with (congestive) heart failure | SNOMED | Heart failure |
| 314378 | 5148006 | Hypertensive heart disease with congestive heart failure | SNOMED | Heart failure |
| 444101 | 46113002 | Hypertensive heart failure | SNOMED | Heart failure |
| 43530961 | 609507007 | Induced termination of pregnancy complicated by cardiac failure | SNOMED | Heart failure |
| 439846 | 85232009 | Left heart failure | SNOMED | Heart failure |
| 4103448 | 25544003 | Low output heart failure | SNOMED | Heart failure |
| 316994 | 83105008 | Malignant hypertensive heart disease with congestive heart failure | SNOMED | Heart failure |
| 46269995 | 11016261000119100 | Nutritional therapy for congestive heart failure done | SNOMED | Heart failure |
| 44782439 | 50781000119109 | Nutrition therapy for congestive heart failure | SNOMED | Heart failure |
| 44811840 | 915571000000102 | On optimal heart failure therapy | SNOMED | Heart failure |
| 36204173 | 86491-8 | Participation in hobbies, recreational activities limited by heart failure over the past 2 weeks [KCCQ] | LOINC | Heart failure |
| 36204176 | 86494-2 | Participation in intimate relationships with loved ones limited by heart failure over the past 2 weeks [KCCQ] | LOINC | Heart failure |
| 36204175 | 86493-4 | Participation in visiting family or friends out of your home limited by heart failure over the past 2 weeks [KCCQ] | LOINC | Heart failure |
| 36204174 | 86492-6 | Participation in working or doing household chores limited by heart failure over the past 2 weeks [KCCQ] | LOINC | Heart failure |
| 4236658 | 90727007 | Pleural effusion due to congestive heart failure | SNOMED | Heart failure |
| 43020910 | 472790001 | Pulmonary hypertension due to left heart disease | SNOMED | Heart failure |
| 4293582 | 403298007 | Red half-moon nail in congestive heart failure | SNOMED | Heart failure |
| 764873 | 7391000175102 | Reduced ejection fraction co-occurrent and due to acute heart failure | SNOMED | Heart failure |
| 764871 | 7371000175103 | Reduced ejection fraction co-occurrent and due to acute on chronic heart failure | SNOMED | Heart failure |
| 764872 | 7381000175100 | Reduced ejection fraction co-occurrent and due to chronic heart failure | SNOMED | Heart failure |
| 4199500 | 314206003 | Refractory heart failure | SNOMED | Heart failure |
| 44809279 | 835981000000108 | Rehabilitation for heart failure | SNOMED | Heart failure |
| 4190773 | 415295002 | Restrictive cardiomyopathy | SNOMED | Heart failure |
| 4184497 | 43736008 | Rheumatic left ventricular failure | SNOMED | Heart failure |
| 4195785 | 44313006 | Right heart failure secondary to left heart failure | SNOMED | Heart failure |
| 4273632 | 367363000 | Right ventricular failure | SNOMED | Heart failure |
| 35615055 | 15964701000119100 | Saddle embolus of pulmonary artery with acute cor pulmonale | SNOMED | Heart failure |
| 44784442 | 698594003 | Symptomatic congestive heart failure | SNOMED | Heart failure |
| 40760358 | 57239-6 | Symptoms in heart failure patients during assessment period [CMS Assessment] | LOINC | Heart failure |
| 443580 | 417996009 | Systolic heart failure | SNOMED | Heart failure |
| 43020421 | 120861000119102 | Systolic heart failure stage C | SNOMED | Heart failure |
| 36712929 | 15629741000119100 | Systolic heart failure stage C due to ischemic cardiomyopathy | SNOMED | Heart failure |
| 43021840 | 120851000119104 | Systolic heart failure stage D | SNOMED | Heart failure |
| 36676642 | 773587008 | X-linked intellectual disability, cardiomegaly, congestive heart failure syndrome | SNOMED | Heart failure |
| 4079695 | 277638005 | Sepsis-associated left ventricular failure | SNOMED | Heart failure |
| 4079296 | 277639002 | Sepsis-associated right ventricular failure | SNOMED | Heart failure |
| 4138307 | 426012001 | Right heart failure due to pulmonary hypertension | SNOMED | Heart failure |
| 4048784 | 230707007 | Anterior cerebral circulation hemorrhagic infarction | SNOMED | Ischemic stroke |
| 4045735 | 230693009 | Anterior cerebral circulation infarction | SNOMED | Ischemic stroke |
| 4031045 | 14309005 | Anterior choroidal artery syndrome | SNOMED | Ischemic stroke |
| 443551 | 428668000 | Apraxia due to cerebrovascular accident | SNOMED | Ischemic stroke |
| 761110 | 151161000119102 | Bilateral cerebral infarction due to precererbral arterial occlusion | SNOMED | Ischemic stroke |
| 4111710 | 195212005 | Brainstem stroke syndrome | SNOMED | Ischemic stroke |
| 4319331 | 95460007 | Cerebellar infarction | SNOMED | Ischemic stroke |
| 4111711 | 195213000 | Cerebellar stroke syndrome | SNOMED | Ischemic stroke |
| 372924 | 20059004 | Cerebral artery occlusion | SNOMED | Ischemic stroke |
| 4110189 | 195185009 | Cerebral infarct due to thrombosis of precerebral arteries | SNOMED | Ischemic stroke |
| 443454 | 432504007 | Cerebral infarction | SNOMED | Ischemic stroke |
| 762951 | 434151000124101 | Cerebral infarction due to anterior cerebral artery occlusion | SNOMED | Ischemic stroke |
| 765515 | 434991000124105 | Cerebral infarction due to basilar artery stenosis | SNOMED | Ischemic stroke |
| 43530683 | 149821000119103 | Cerebral infarction due to carotid artery occlusion | SNOMED | Ischemic stroke |
| 762933 | 433891000124100 | Cerebral infarction due to cerebral artery occlusion | SNOMED | Ischemic stroke |
| 762937 | 433961000124100 | Cerebral infarction due to cerebral venous thrombosis | SNOMED | Ischemic stroke |
| 4111714 | 195230003 | Cerebral infarction due to cerebral venous thrombosis, non-pyogenic | SNOMED | Ischemic stroke |
| 4108356 | 195190007 | Cerebral infarction due to embolism of cerebral arteries | SNOMED | Ischemic stroke |
| 45772786 | 705128004 | Cerebral infarction due to embolism of middle cerebral artery | SNOMED | Ischemic stroke |
| 4110190 | 195186005 | Cerebral infarction due to embolism of precerebral arteries | SNOMED | Ischemic stroke |
| 762935 | 433931000124109 | Cerebral infarction due to internal carotid artery occlusion | SNOMED | Ischemic stroke |
| 763015 | 434961000124102 | Cerebral infarction due to middle cerebral artery occlusion | SNOMED | Ischemic stroke |
| 46273649 | 34181000119102 | Cerebral infarction due to occlusion of basilar artery | SNOMED | Ischemic stroke |
| 35610084 | 1089411000000100 | Cerebral infarction due to occlusion of cerebral artery | SNOMED | Ischemic stroke |
| 46270031 | 125081000119106 | Cerebral infarction due to occlusion of precerebral artery | SNOMED | Ischemic stroke |
| 762934 | 433911000124103 | Cerebral infarction due to posterior cerebral artery occlusion | SNOMED | Ischemic stroke |
| 43531607 | 99451000119105 | Cerebral infarction due to stenosis of carotid artery | SNOMED | Ischemic stroke |
| 35610085 | 1089421000000100 | Cerebral infarction due to stenosis of cerebral artery | SNOMED | Ischemic stroke |
| 46270381 | 293831000119105 | Cerebral infarction due to stenosis of precerebral artery | SNOMED | Ischemic stroke |
| 4110192 | 195189003 | Cerebral infarction due to thrombosis of cerebral arteries | SNOMED | Ischemic stroke |
| 45767658 | 705130002 | Cerebral infarction due to thrombosis of middle cerebral artery | SNOMED | Ischemic stroke |
| 44782773 | 34191000119104 | Cerebral infarction due to vertebral artery occlusion | SNOMED | Ischemic stroke |
| 46270380 | 293811000119100 | Cerebral infarction due to vertebral artery stenosis | SNOMED | Ischemic stroke |
| 37110678 | 724993002 | Cerebral ischemic stroke due to occlusion of extracranial large artery | SNOMED | Ischemic stroke |
| 37110679 | 724994008 | Cerebral ischemic stroke due to stenosis of extracranial large artery | SNOMED | Ischemic stroke |
| 381316 | 230690007 | Cerebrovascular accident | SNOMED | Ischemic stroke |
| 4045734 | 230691006 | CVA - cerebrovascular accident due to cerebral artery occlusion | SNOMED | Ischemic stroke |
| 443465 | 426033005 | Dysphagia as a late effect of cerebrovascular accident | SNOMED | Ischemic stroke |
| 44782781 | 48601000119107 | Hemiplegia and/or hemiparesis following stroke | SNOMED | Ischemic stroke |
| 4046362 | 230706003 | Hemorrhagic cerebral infarction | SNOMED | Ischemic stroke |
| 4131383 | 413102000 | Infarction of basal ganglia | SNOMED | Ischemic stroke |
| 4046237 | 230523009 | Infarction of optic radiation | SNOMED | Ischemic stroke |
| 4119140 | 302904002 | Infarction of visual cortex | SNOMED | Ischemic stroke |
| 4043731 | 230692004 | Infarction - precerebral | SNOMED | Ischemic stroke |
| 4046360 | 230698000 | Lacunar infarction | SNOMED | Ischemic stroke |
| 4141405 | 307766002 | Left sided cerebral infarction | SNOMED | Ischemic stroke |
| 4110194 | 195209007 | Middle cerebral artery syndrome | SNOMED | Ischemic stroke |
| 197303 | 425642008 | Monoplegia of dominant lower limb as a late effect of cerebrovascular accident | SNOMED | Ischemic stroke |
| 443525 | 427065003 | Monoplegia of dominant upper limb as a late effect of cerebrovascular accident | SNOMED | Ischemic stroke |
| 40480946 | 441894009 | Monoplegia of nondominant lower limb as a late effect of cerebrovascular accident | SNOMED | Ischemic stroke |
| 37116473 | 733199002 | Multifocal cerebral infarction due to and following procedure on cardiovascular system | SNOMED | Ischemic stroke |
| 4077086 | 276219001 | Occipital cerebral infarction | SNOMED | Ischemic stroke |
| 372654 | 425882004 | Paralytic syndrome as late effect of stroke | SNOMED | Ischemic stroke |
| 443609 | 430959006 | Paralytic syndrome of dominant side as late effect of stroke | SNOMED | Ischemic stroke |
| 443599 | 430947007 | Paralytic syndrome of nondominant side as late effect of stroke | SNOMED | Ischemic stroke |
| 43530742 | 361000119103 | Paralytic syndrome on one side of the body as late effect of cerebrovascular accident | SNOMED | Ischemic stroke |
| 4046359 | 230695002 | Partial anterior cerebral circulation infarction | SNOMED | Ischemic stroke |
| 4319146 | 95830009 | Pituitary infarction | SNOMED | Ischemic stroke |
| 4043732 | 230708002 | Posterior cerebral circulation hemorrhagic infarction | SNOMED | Ischemic stroke |
| 4045737 | 230699008 | Pure motor lacunar infarction | SNOMED | Ischemic stroke |
| 4045738 | 230700009 | Pure sensory lacunar infarction | SNOMED | Ischemic stroke |
| 40482301 | 442212003 | Residual cognitive deficit as late effect of cerebrovascular accident | SNOMED | Ischemic stroke |
| 4146185 | 307767006 | Right sided cerebral infarction | SNOMED | Ischemic stroke |
| 4112026 | 195243003 | Sequelae of cerebral infarction | SNOMED | Ischemic stroke |
| 36717605 | 723082006 | Silent cerebral infarct | SNOMED | Ischemic stroke |
| 40481354 | 441960006 | Speech and language deficit as late effect of cerebrovascular accident | SNOMED | Ischemic stroke |
| 4142739 | 427296003 | Thalamic infarction | SNOMED | Ischemic stroke |
| 4046358 | 230694003 | Total anterior cerebral circulation infarction | SNOMED | Ischemic stroke |
| 43530744 | 40161000119102 | Weakness of face muscles as sequela of stroke | SNOMED | Ischemic stroke |
| 40479572 | 441526008 | Infarct of cerebrum due to iatrogenic cerebrovascular accident | SNOMED | Ischemic stroke |
| 4120091 | 233936003 | Acute massive pulmonary embolism | SNOMED | Pulmonary embolism (PE) |
| 45768439 | 706870000 | Acute pulmonary embolism | SNOMED | Pulmonary embolism (PE) |
| 45768888 | 707414004 | Acute pulmonary thromboembolism | SNOMED | Pulmonary embolism (PE) |
| 762808 | 432201000124103 | Infarction of lung due to embolus | SNOMED | Pulmonary embolism (PE) |
| 40480461 | 441746000 | Infarction of lung due to iatrogenic pulmonary embolism | SNOMED | Pulmonary embolism (PE) |
| 440417 | 59282003 | Pulmonary embolism | SNOMED | Pulmonary embolism (PE) |
| 37016922 | 713078005 | Pulmonary embolism on long-term anticoagulation therapy | SNOMED | Pulmonary embolism (PE) |
| 40761384 | 58283-3 | Pulmonary embolism requiring hospitalization | LOINC | Pulmonary embolism (PE) |
| 43530605 | 1001000119102 | Pulmonary embolism with pulmonary infarction | SNOMED | Pulmonary embolism (PE) |
| 4253796 | 74315008 | Pulmonary microemboli | SNOMED | Pulmonary embolism (PE) |
| 4121618 | 233935004 | Pulmonary thromboembolism | SNOMED | Pulmonary embolism (PE) |
| 36713113 | 328511000119109 | Saddle embolus of pulmonary artery | SNOMED | Pulmonary embolism (PE) |
| 35615055 | 15964701000119100 | Saddle embolus of pulmonary artery with acute cor pulmonale | SNOMED | Pulmonary embolism (PE) |
| 4119607 | 233937007 | Subacute massive pulmonary embolism | SNOMED | Pulmonary embolism (PE) |
| 4027729 | 12847006 | Acute duodenal ulcer with hemorrhage | SNOMED | GI hemorrhage |
| 441062 | 87756006 | Acute duodenal ulcer with hemorrhage AND obstruction | SNOMED | GI hemorrhage |
| 4336230 | 86895006 | Acute duodenal ulcer with hemorrhage AND perforation | SNOMED | GI hemorrhage |
| 435855 | 51847008 | Acute duodenal ulcer with hemorrhage AND with perforation but without obstruction | SNOMED | GI hemorrhage |
| 434402 | 66767006 | Acute duodenal ulcer with hemorrhage but without obstruction | SNOMED | GI hemorrhage |
| 437021 | 41986000 | Acute duodenal ulcer with hemorrhage, with perforation AND with obstruction | SNOMED | GI hemorrhage |
| 4231580 | 89748001 | Acute gastric ulcer with hemorrhage | SNOMED | GI hemorrhage |
| 198467 | 46708007 | Acute gastric ulcer with hemorrhage and obstruction | SNOMED | GI hemorrhage |
| 4169592 | 48974009 | Acute gastric ulcer with hemorrhage and perforation | SNOMED | GI hemorrhage |
| 199855 | 17067009 | Acute gastric ulcer with hemorrhage AND with perforation but without obstruction | SNOMED | GI hemorrhage |
| 193795 | 70418001 | Acute gastric ulcer with hemorrhage but without obstruction | SNOMED | GI hemorrhage |
| 195845 | 53337006 | Acute gastric ulcer with hemorrhage, with perforation and with obstruction | SNOMED | GI hemorrhage |
| 4100660 | 27719009 | Acute gastrointestinal hemorrhage | SNOMED | GI hemorrhage |
| 4274491 | 63954007 | Acute gastrojejunal ulcer with hemorrhage | SNOMED | GI hemorrhage |
| 441063 | 72408002 | Acute gastrojejunal ulcer with hemorrhage and obstruction | SNOMED | GI hemorrhage |
| 4217947 | 81387001 | Acute gastrojejunal ulcer with hemorrhage and perforation | SNOMED | GI hemorrhage |
| 441328 | 66673003 | Acute gastrojejunal ulcer with hemorrhage and with perforation but without obstruction | SNOMED | GI hemorrhage |
| 438468 | 59515005 | Acute gastrojejunal ulcer with hemorrhage but without obstruction | SNOMED | GI hemorrhage |
| 442314 | 58711008 | Acute gastrojejunal ulcer with hemorrhage, with perforation and with obstruction | SNOMED | GI hemorrhage |
| 4320492 | 69810009 | Acute hemorrhagic colitis caused by Escherichia coli | SNOMED | GI hemorrhage |
| 4340509 | 236016008 | Acute hemorrhagic enterocolitis | SNOMED | GI hemorrhage |
| 193249 | 2367005 | Acute hemorrhagic gastritis | SNOMED | GI hemorrhage |
| 36717494 | 721690003 | Acute hemorrhagic ulcer of rectum | SNOMED | GI hemorrhage |
| 4310668 | 85521005 | Acute lower gastrointestinal hemorrhage | SNOMED | GI hemorrhage |
| 4046500 | 12274003 | Acute peptic ulcer with hemorrhage | SNOMED | GI hemorrhage |
| 23237 | 43406003 | Acute peptic ulcer with hemorrhage AND obstruction | SNOMED | GI hemorrhage |
| 4006994 | 111353003 | Acute peptic ulcer with hemorrhage and perforation | SNOMED | GI hemorrhage |
| 27026 | 47064007 | Acute peptic ulcer with hemorrhage AND with perforation but without obstruction | SNOMED | GI hemorrhage |
| 31335 | 22157005 | Acute peptic ulcer with hemorrhage but without obstruction | SNOMED | GI hemorrhage |
| 194986 | 28945005 | Acute peptic ulcer with hemorrhage, with perforation AND with obstruction | SNOMED | GI hemorrhage |
| 4308202 | 38938002 | Acute upper gastrointestinal hemorrhage | SNOMED | GI hemorrhage |
| 4173032 | 276465001 | Anal margin hematoma | SNOMED | GI hemorrhage |
| 3171162 | 6210001000004100 | Angiodysplasia of intestine with hemorrhage | Nebraska Lexicon | GI hemorrhage |
| 4095572 | 262858002 | Appendix hematoma | SNOMED | GI hemorrhage |
| 1077328 | 46751000087108 | Benign neoplasm of large intestine with hemorrhage | SNOMED | GI hemorrhage |
| 4096916 | 26421009 | Bleeding external hemorrhoids | SNOMED | GI hemorrhage |
| 4245614 | 6072007 | Bleeding from anus | SNOMED | GI hemorrhage |
| 3175732 | 13590001000004100 | Bleeding from colostomy | Nebraska Lexicon | GI hemorrhage |
| 4087310 | 24807004 | Bleeding gastric varices | SNOMED | GI hemorrhage |
| 4198840 | 51551000 | Bleeding hemorrhoids | SNOMED | GI hemorrhage |
| 608969 | 15970261000119100 | Bleeding internal hemorrhoid grade I | SNOMED | GI hemorrhage |
| 608970 | 15970301000119100 | Bleeding internal hemorrhoid grade II | SNOMED | GI hemorrhage |
| 608968 | 15970021000119100 | Bleeding internal hemorrhoid grade III | SNOMED | GI hemorrhage |
| 608971 | 15970341000119100 | Bleeding internal hemorrhoid grade IV | SNOMED | GI hemorrhage |
| 4295428 | 75884004 | Bleeding internal hemorrhoids | SNOMED | GI hemorrhage |
| 40482450 | 445162006 | Bleeding Meckel's diverticulitis | SNOMED | GI hemorrhage |
| 40482451 | 445163001 | Bleeding Meckel's diverticulum | SNOMED | GI hemorrhage |
| 4205670 | 308882008 | Bleeding stress ulcer of stomach | SNOMED | GI hemorrhage |
| 26441 | 57748001 | Bleeding ulcer of esophagus | SNOMED | GI hemorrhage |
| 1077329 | 46761000087106 | Bleeding ulcer of large intestine | SNOMED | GI hemorrhage |
| 1077330 | 46771000087102 | Bleeding ulcer of mucosa of large intestine | SNOMED | GI hemorrhage |
| 4119169 | 262865005 | Cecal hematoma | SNOMED | GI hemorrhage |
| 44784282 | 698352000 | Chronic antral gastritis with hemorrhage | SNOMED | GI hemorrhage |
| 4232181 | 89469000 | Chronic duodenal ulcer with hemorrhage | SNOMED | GI hemorrhage |
| 437323 | 34021006 | Chronic duodenal ulcer with hemorrhage AND obstruction | SNOMED | GI hemorrhage |
| 4289830 | 36975000 | Chronic duodenal ulcer with hemorrhage AND perforation | SNOMED | GI hemorrhage |
| 438796 | 81142005 | Chronic duodenal ulcer with hemorrhage AND with perforation but without obstruction | SNOMED | GI hemorrhage |
| 436148 | 62341002 | Chronic duodenal ulcer with hemorrhage but without obstruction | SNOMED | GI hemorrhage |
| 440756 | 86258000 | Chronic duodenal ulcer with hemorrhage, with perforation AND with obstruction | SNOMED | GI hemorrhage |
| 4211001 | 57246001 | Chronic gastric ulcer with hemorrhage | SNOMED | GI hemorrhage |
| 201885 | 85859006 | Chronic gastric ulcer with hemorrhage and with obstruction | SNOMED | GI hemorrhage |
| 4294973 | 76181002 | Chronic gastric ulcer with hemorrhage and with perforation | SNOMED | GI hemorrhage |
| 196442 | 74341002 | Chronic gastric ulcer with hemorrhage AND with perforation but without obstruction | SNOMED | GI hemorrhage |
| 197018 | 76078009 | Chronic gastric ulcer with hemorrhage but without obstruction | SNOMED | GI hemorrhage |
| 198801 | 85787009 | Chronic gastric ulcer with hemorrhage, with perforation and with obstruction | SNOMED | GI hemorrhage |
| 4307661 | 83628005 | Chronic gastrointestinal hemorrhage | SNOMED | GI hemorrhage |
| 433515 | 62838000 | Chronic gastrojejunal ulcer with hemorrhage | SNOMED | GI hemorrhage |
| 436729 | 90257004 | Chronic gastrojejunal ulcer with hemorrhage and obstruction | SNOMED | GI hemorrhage |
| 4164920 | 45640006 | Chronic gastrojejunal ulcer with hemorrhage and perforation | SNOMED | GI hemorrhage |
| 437326 | 46523000 | Chronic gastrojejunal ulcer with hemorrhage and with perforation but without obstruction | SNOMED | GI hemorrhage |
| 4244406 | 59356009 | Chronic gastrojejunal ulcer with hemorrhage but without perforation | SNOMED | GI hemorrhage |
| 443779 | 24001002 | Chronic gastrojejunal ulcer with hemorrhage, with perforation and with obstruction | SNOMED | GI hemorrhage |
| 4195068 | 68162000 | Chronic lower gastrointestinal hemorrhage | SNOMED | GI hemorrhage |
| 4174044 | 49232000 | Chronic peptic ulcer with hemorrhage | SNOMED | GI hemorrhage |
| 24076 | 56461008 | Chronic peptic ulcer with hemorrhage AND obstruction | SNOMED | GI hemorrhage |
| 4247008 | 61300005 | Chronic peptic ulcer with hemorrhage AND perforation | SNOMED | GI hemorrhage |
| 22665 | 55746001 | Chronic peptic ulcer with hemorrhage AND with perforation but without obstruction | SNOMED | GI hemorrhage |
| 30770 | 81518000 | Chronic peptic ulcer with hemorrhage but without obstruction | SNOMED | GI hemorrhage |
| 24397 | 77661009 | Chronic peptic ulcer with hemorrhage, with perforation AND with obstruction | SNOMED | GI hemorrhage |
| 4103011 | 25349007 | Chronic upper gastrointestinal hemorrhage | SNOMED | GI hemorrhage |
| 4246979 | 40835002 | Coffee ground vomiting | SNOMED | GI hemorrhage |
| 4095094 | 262870003 | Colonic hematoma | SNOMED | GI hemorrhage |
| 3171798 | 5600001000004100 | Diverticulitis of colon with hemorrhage | Nebraska Lexicon | GI hemorrhage |
| 36685122 | 1086591000119100 | Diverticulosis of sigmoid colon with hemorrhage | SNOMED | GI hemorrhage |
| 42536738 | 735729001 | Diverticulum and hemorrhage of large intestine | SNOMED | GI hemorrhage |
| 42536736 | 735727004 | Diverticulum and hemorrhage of small intestine | SNOMED | GI hemorrhage |
| 4342987 | 236095006 | Duodenal anastomotic hemorrhage | SNOMED | GI hemorrhage |
| 4096032 | 262843005 | Duodenal hematoma | SNOMED | GI hemorrhage |
| 4318535 | 95533003 | Duodenal hemorrhage | SNOMED | GI hemorrhage |
| 46269821 | 1082701000119100 | Duodenal hemorrhage due to angiodysplasia of duodenum | SNOMED | GI hemorrhage |
| 46269892 | 1086031000119100 | Duodenal hemorrhage due to Dieulafoy lesion of duodenum | SNOMED | GI hemorrhage |
| 4099014 | 27281001 | Duodenal ulcer with hemorrhage | SNOMED | GI hemorrhage |
| 4084844 | 18367003 | Duodenal ulcer with hemorrhage AND obstruction | SNOMED | GI hemorrhage |
| 4031954 | 23812009 | Duodenal ulcer with hemorrhage AND perforation | SNOMED | GI hemorrhage |
| 4035167 | 15115006 | Duodenal ulcer with hemorrhage AND with perforation but without obstruction | SNOMED | GI hemorrhage |
| 4149010 | 35560008 | Duodenal ulcer with hemorrhage but without obstruction | SNOMED | GI hemorrhage |
| 4049270 | 12355008 | Duodenal ulcer with hemorrhage, with perforation AND with obstruction | SNOMED | GI hemorrhage |
| 606751 | 1148555003 | Erosion of duodenum with hemorrhage | SNOMED | GI hemorrhage |
| 4143871 | 307233002 | Erosion of stomach with hemorrhage | SNOMED | GI hemorrhage |
| 4341252 | 236093004 | Esophageal anastomotic hemorrhage | SNOMED | GI hemorrhage |
| 23245 | 15238002 | Esophageal bleeding | SNOMED | GI hemorrhage |
| 4232237 | 439442003 | Esophageal bleeding due to ulcerative esophagitis | SNOMED | GI hemorrhage |
| 4095555 | 262790002 | Esophageal hematoma | SNOMED | GI hemorrhage |
| 1340239 | OMOP5165894 | Exacerbation of acute hemorrhagic gastritis | OMOP Extension | GI hemorrhage |
| 4342905 | 236094005 | Gastric anastomotic hemorrhage | SNOMED | GI hemorrhage |
| 193250 | 61401005 | Gastric hemorrhage | SNOMED | GI hemorrhage |
| 46269910 | 1087081000119100 | Gastric hemorrhage caused by Helicobacter pylori | SNOMED | GI hemorrhage |
| 45757783 | 40241000119109 | Gastric hemorrhage due to alcoholic gastritis | SNOMED | GI hemorrhage |
| 46269819 | 1082631000119100 | Gastric hemorrhage due to allergic gastritis | SNOMED | GI hemorrhage |
| 46269822 | 1082711000119100 | Gastric hemorrhage due to angiodysplasia of stomach | SNOMED | GI hemorrhage |
| 46270145 | 150721000119102 | Gastric hemorrhage due to atrophic gastritis | SNOMED | GI hemorrhage |
| 46269837 | 1085121000119100 | Gastric hemorrhage due to chronic superficial gastritis | SNOMED | GI hemorrhage |
| 46269893 | 1086041000119100 | Gastric hemorrhage due to Dieulafoy lesion of stomach | SNOMED | GI hemorrhage |
| 46270025 | 123411000119106 | Gastric hemorrhage due to eosinophilic gastritis | SNOMED | GI hemorrhage |
| 45768629 | 7071000119102 | Gastric hemorrhage due to erosive gastritis | SNOMED | GI hemorrhage |
| 46269911 | 1087161000119100 | Gastric hemorrhage due to hypertrophic gastritis | SNOMED | GI hemorrhage |
| 45757570 | 218541000119109 | Gastric hemorrhage due to idiopathic erosive gastritis | SNOMED | GI hemorrhage |
| 46269912 | 1087811000119100 | Gastric hemorrhage due to irritant gastritis | SNOMED | GI hemorrhage |
| 46269935 | 1090361000119100 | Gastric hemorrhage due to pyloric gastritis | SNOMED | GI hemorrhage |
| 46269908 | 1086951000119100 | Gastric hemorrhage due to vascular ectasia of gastric antrum | SNOMED | GI hemorrhage |
| 46269953 | 1092941000119100 | Gastric hemorrhage due to viral gastritis | SNOMED | GI hemorrhage |
| 4049466 | 15902003 | Gastric ulcer with hemorrhage | SNOMED | GI hemorrhage |
| 4207217 | 53877005 | Gastric ulcer with hemorrhage and obstruction | SNOMED | GI hemorrhage |
| 4266523 | 62366003 | Gastric ulcer with hemorrhage and perforation | SNOMED | GI hemorrhage |
| 4071203 | 2066005 | Gastric ulcer with hemorrhage AND perforation but without obstruction | SNOMED | GI hemorrhage |
| 4041707 | 16694003 | Gastric ulcer with hemorrhage but without obstruction | SNOMED | GI hemorrhage |
| 4069838 | 17593008 | Gastric ulcer with hemorrhage, with perforation and with obstruction | SNOMED | GI hemorrhage |
| 36684448 | 97801000119102 | Gastritis with upper gastrointestinal hemorrhage | SNOMED | GI hemorrhage |
| 4342904 | 236092009 | Gastrointestinal anastomotic hemorrhage | SNOMED | GI hemorrhage |
| 192671 | 74474003 | Gastrointestinal hemorrhage | SNOMED | GI hemorrhage |
| 37163132 | 1204464003 | Gastrointestinal hemorrhage with sepsis | SNOMED | GI hemorrhage |
| 4222477 | 84124004 | Gastrojejunal ulcer with hemorrhage | SNOMED | GI hemorrhage |
| 4175673 | 42698006 | Gastrojejunal ulcer with hemorrhage AND obstruction | SNOMED | GI hemorrhage |
| 4273874 | 64094003 | Gastrojejunal ulcer with hemorrhage AND perforation | SNOMED | GI hemorrhage |
| 4336971 | 87796008 | Gastrojejunal ulcer with hemorrhage and with perforation but without obstruction | SNOMED | GI hemorrhage |
| 4179773 | 50663005 | Gastrojejunal ulcer with hemorrhage but without obstruction | SNOMED | GI hemorrhage |
| 4183005 | 54798007 | Gastrojejunal ulcer with hemorrhage, with perforation and with obstruction | SNOMED | GI hemorrhage |
| 26727 | 8765009 | Hematemesis | SNOMED | GI hemorrhage |
| 4204041 | 308904008 | Hematemesis - cause unknown | SNOMED | GI hemorrhage |
| 4085415 | 282074002 | Hematoma of anus | SNOMED | GI hemorrhage |
| 4106997 | 282065004 | Hematoma of ileum | SNOMED | GI hemorrhage |
| 4083969 | 282054009 | Hematoma of jejunum | SNOMED | GI hemorrhage |
| 46272978 | 711441008 | Hemorrhage of cecum | SNOMED | GI hemorrhage |
| 46273470 | 1086551000119100 | Hemorrhage of cecum due to diverticulosis | SNOMED | GI hemorrhage |
| 442190 | 95540002 | Hemorrhage of colon | SNOMED | GI hemorrhage |
| 37161060 | 1173099009 | Hemorrhage of colon due to angiodysplasia of colon | SNOMED | GI hemorrhage |
| 45757543 | 190191000119107 | Hemorrhage of colon due to diverticulosis | SNOMED | GI hemorrhage |
| 37174298 | 805616441000119000 | Hemorrhage of ileostomy stoma | SNOMED | GI hemorrhage |
| 46273182 | 712509002 | Hemorrhage of jejunum | SNOMED | GI hemorrhage |
| 46269906 | 1086581000119100 | Hemorrhage of jejunum with diverticulosis | SNOMED | GI hemorrhage |
| 37162897 | 1197741006 | Hemorrhage of large intestine due to diverticulitis | SNOMED | GI hemorrhage |
| 194395 | 197092000 | Hemorrhage of large intestine with diverticular disease of large intestine | SNOMED | GI hemorrhage |
| 37162899 | 1197743009 | Hemorrhage of lower esophagus due to erosive gastro-esophageal reflux disease | SNOMED | GI hemorrhage |
| 197925 | 266464001 | Hemorrhage of rectum and anus | SNOMED | GI hemorrhage |
| 46269901 | 1086461000119100 | Hemorrhage of small intestine due to diverticulitis | SNOMED | GI hemorrhage |
| 46270529 | 40271000119102 | Hemorrhage of small intestine with diverticulosis | SNOMED | GI hemorrhage |
| 46269904 | 1086561000119100 | Hemorrhage with diverticulosis of duodenum | SNOMED | GI hemorrhage |
| 46269905 | 1086571000119100 | Hemorrhage with diverticulosis of ileum | SNOMED | GI hemorrhage |
| 4216673 | 81318004 | Hemorrhagic colitis | SNOMED | GI hemorrhage |
| 437027 | 95531001 | Hemorrhagic duodenitis | SNOMED | GI hemorrhage |
| 4134808 | 413212000 | Hemorrhagic duodenopathy | SNOMED | GI hemorrhage |
| 4128705 | 235224000 | Hemorrhagic enteritis of intestine | SNOMED | GI hemorrhage |
| 4048889 | 15720001 | Hemorrhagic enteropathy of terminal ileum | SNOMED | GI hemorrhage |
| 4264487 | 60698006 | Hemorrhagic esophagitis | SNOMED | GI hemorrhage |
| 4260059 | 409506009 | Hemorrhagic gastroenteritis | SNOMED | GI hemorrhage |
| 4131525 | 413218001 | Hemorrhagic gastropathy | SNOMED | GI hemorrhage |
| 4188154 | 414390000 | Hemorrhagic mucosa of duodenum | SNOMED | GI hemorrhage |
| 4185784 | 414391001 | Hemorrhagic mucosa of stomach | SNOMED | GI hemorrhage |
| 4322061 | 981008 | Hemorrhagic proctitis | SNOMED | GI hemorrhage |
| 4318829 | 95535005 | Ileal hemorrhage | SNOMED | GI hemorrhage |
| 46273183 | 712510007 | Intestinal hemorrhage | SNOMED | GI hemorrhage |
| 45757654 | 29731000119103 | Intestinal hemorrhage due to angiodysplasia of intestine | SNOMED | GI hemorrhage |
| 46269907 | 1086601000119100 | Intestinal hemorrhage with diverticulosis | SNOMED | GI hemorrhage |
| 4050203 | 15970005 | Intramural esophageal hematoma | SNOMED | GI hemorrhage |
| 36717644 | 717882009 | Intramural hemorrhage of duodenum | SNOMED | GI hemorrhage |
| 36715487 | 721179009 | Intramural hemorrhage of esophagus | SNOMED | GI hemorrhage |
| 36713507 | 717868009 | Intramural hemorrhage of stomach | SNOMED | GI hemorrhage |
| 37017911 | 714278008 | Jejunal anastomotic hemorrhage | SNOMED | GI hemorrhage |
| 4341790 | 236097003 | Large intestine anastomotic hemorrhage | SNOMED | GI hemorrhage |
| 4338544 | 87763006 | Lower gastrointestinal hemorrhage | SNOMED | GI hemorrhage |
| 1077332 | 46811000087102 | Malignant neoplasm of large intestine with hemorrhage | SNOMED | GI hemorrhage |
| 316457 | 35265002 | Mallory-Weiss syndrome | SNOMED | GI hemorrhage |
| 4139411 | 307297004 | Massive gastrointestinal bleed | SNOMED | GI hemorrhage |
| 37109016 | 16055151000119100 | Neonatal gastrointestinal hemorrhage | SNOMED | GI hemorrhage |
| 4071070 | 206423004 | Neonatal hematemesis | SNOMED | GI hemorrhage |
| 4048286 | 206425006 | Neonatal rectal hemorrhage | SNOMED | GI hemorrhage |
| 36717237 | 722886001 | Obscure gastrointestinal hemorrhage | SNOMED | GI hemorrhage |
| 4271696 | 64121000 | Peptic ulcer with hemorrhage | SNOMED | GI hemorrhage |
| 4273101 | 64398008 | Peptic ulcer with hemorrhage AND obstruction | SNOMED | GI hemorrhage |
| 4206466 | 55617001 | Peptic ulcer with hemorrhage AND perforation | SNOMED | GI hemorrhage |
| 4235740 | 90489006 | Peptic ulcer with hemorrhage AND with perforation but without obstruction | SNOMED | GI hemorrhage |
| 4169394 | 48658001 | Peptic ulcer with hemorrhage but without obstruction | SNOMED | GI hemorrhage |
| 4095410 | 26221006 | Peptic ulcer with hemorrhage, with perforation AND with obstruction | SNOMED | GI hemorrhage |
| 46269902 | 1086481000119100 | Perforation of small intestine co-occurrent with hemorrhage due to diverticulitis | SNOMED | GI hemorrhage |
| 194158 | 48729005 | Perinatal gastrointestinal hemorrhage | SNOMED | GI hemorrhage |
| 4048601 | 206406008 | Perinatal hematemesis | SNOMED | GI hemorrhage |
| 4048602 | 206408009 | Perinatal rectal hemorrhage | SNOMED | GI hemorrhage |
| 46271323 | 709424004 | Postoperative hemorrhage of anus | SNOMED | GI hemorrhage |
| 37160869 | 1163589006 | Prepyloric ulcer with hemorrhage | SNOMED | GI hemorrhage |
| 4096781 | 262876009 | Rectal hematoma | SNOMED | GI hemorrhage |
| 4026112 | 12063002 | Rectal hemorrhage | SNOMED | GI hemorrhage |
| 1075403 | 1303925000 | Rectal hemorrhage caused by apixaban | SNOMED | GI hemorrhage |
| 1075402 | 1303924001 | Rectal hemorrhage caused by drug | SNOMED | GI hemorrhage |
| 46269841 | 1085171000119100 | Rectal hemorrhage due to chronic ulcerative pancolitis | SNOMED | GI hemorrhage |
| 46269847 | 1085221000119100 | Rectal hemorrhage due to chronic ulcerative proctitis | SNOMED | GI hemorrhage |
| 46269851 | 1085271000119100 | Rectal hemorrhage due to chronic ulcerative rectosigmoiditis | SNOMED | GI hemorrhage |
| 46269891 | 1085941000119100 | Rectal hemorrhage due to Crohn's disease | SNOMED | GI hemorrhage |
| 46269877 | 1085791000119100 | Rectal hemorrhage due to Crohn's disease of large intestine | SNOMED | GI hemorrhage |
| 46269882 | 1085841000119100 | Rectal hemorrhage due to Crohn's disease of small and large intestines | SNOMED | GI hemorrhage |
| 46269887 | 1085891000119100 | Rectal hemorrhage due to Crohn's disease of small intestine | SNOMED | GI hemorrhage |
| 46269863 | 1085431000119100 | Rectal hemorrhage due to inflammatory polyps of colon | SNOMED | GI hemorrhage |
| 46273478 | 1092881000119100 | Rectal hemorrhage due to ulcerative colitis | SNOMED | GI hemorrhage |
| 4144926 | 307296008 | Recurrent gastrointestinal bleeding | SNOMED | GI hemorrhage |
| 4095570 | 262852001 | Small intestinal hematoma | SNOMED | GI hemorrhage |
| 197024 | 70375006 | Small intestinal hemorrhage | SNOMED | GI hemorrhage |
| 4341253 | 236096007 | Small intestine anastomotic hemorrhage | SNOMED | GI hemorrhage |
| 4096773 | 262838003 | Stomach hematoma | SNOMED | GI hemorrhage |
| 4291649 | 37372002 | Upper gastrointestinal bleeding | SNOMED | GI hemorrhage |
| 4332645 | 430349003 | Upper gastrointestinal hemorrhage associated with hypercoagulability state | SNOMED | GI hemorrhage |
| 3175463 | 8440001000004100 | Vascular ectasia of stomach and duodenum with hemorrhage | Nebraska Lexicon | GI hemorrhage |
| 4145805 | 267051003 | Vomiting blood - fresh | SNOMED | GI hemorrhage |
| 607946 | 1156385007 | Wischnewski spots | SNOMED | GI hemorrhage |

### Table S3: Baseline characteristics of individuals with record of individual NSAID prescriptions and cardiovascular events of interest (excluding aspirin). Patients characterised on index date of first NSAID-CV event combination window, which would either be the first day of NSAID prescription or date of CV event. Only those with more than 50 records included.

| **Description** | **Celecoxib** | **Etoricoxib** | **Diclofenac** | **Etodolac** | **Ibuprofen** | **Indomethacin** | **Mefenamate** | **Meloxicam** | **Naproxen** |
| --- | --- | --- | --- | --- | --- | --- | --- | --- | --- |
| Number subjects | 346 | 480 | 1,718 | 96 | 5,980 | 300 | 178 | 419 | 9,773 |
| Age (Median [Q25 - Q75]) | 68 [57 - 77] | 70 [57 - 79] | 62 [50 - 72] | 71 [62 - 78] | 68 [56 - 78] | 71 [62 - 78] | 46 [38 - 52] | 71 [60 - 79] | 65 [54 - 76] |
| **Age group** |  |  |  |  |  |  |  |  |  |
| 18 to 49 | 46 (13.3%) | 58 (12.1%) | 409 (23.8%) | 5 (5.2%) | 992 (16.6%) | 22 (7.3%) | 117 (65.7%) | 36 (8.6%) | 1,723 (17.6%) |
| 50 to 59 | 58 (16.8%) | 78 (16.3%) | 369 (21.5%) | 15 (15.6%) | 919 (15.4%) | 38 (12.7%) | 30 (16.9%) | 65 (15.5%) | 1,814 (18.6%) |
| 60 to 69 | 81 (23.4%) | 88 (18.3%) | 418 (24.3%) | 25 (26.0%) | 1,312 (21.9%) | 74 (24.7%) | 12 (6.7%) | 90 (21.5%) | 2,309 (23.6%) |
| 70 to 79 | 96 (27.8%) | 156 (32.5%) | 358 (20.8%) | 31 (32.3%) | 1,452 (24.3%) | 100 (33.3%) | 9 (5.1%) | 130 (31.0%) | 2,381 (24.4%) |
| 80 to 89 | 56 (16.2%) | 81 (16.9%) | 143 (8.3%) | 18 (18.8%) | 1,067 (17.8%) | 60 (20.0%) | 10 (5.6%) | 81 (19.3%) | 1,337 (13.7%) |
| 90+ | 9 (2.6%) | 19 (4.0%) | 21 (1.2%) | <5 | 238 (4.0%) | 6 (2.0%) | - | 17 (4.1%) | 209 (2.1%) |
| **Sex** |  |  |  |  |  |  |  |  |  |
| Male | 147 (42.5%) | 263 (54.8%) | 956 (55.7%) | 44 (45.8%) | 3,080 (51.5%) | 214 (71.3%) | 11 (6.2%) | 190 (45.4%) | 5,516 (56.4%) |
| Female | 199 (57.5%) | 217 (45.2%) | 762 (44.4%) | 52 (54.2%) | 2,900 (48.5%) | 86 (28.7%) | 167 (93.8%) | 229 (54.7%) | 4,257 (43.6%) |
| **Conditions prior to index date (N %)** |  |  |  |  |  |  |  |  |  |
| Uti | 53 (15.3%) | 65 (13.5%) | 193 (11.2%) | 12 (12.5%) | 854 (14.3%) | 41 (13.7%) | 25 (14.0%) | 63 (15.0%) | 1,268 (13.0%) |
| Cerebrovascular disease | 19 (5.5%) | 24 (5.0%) | 67 (3.9%) | <5 | 360 (6.0%) | 24 (8.0%) | <5 | 35 (8.4%) | 518 (5.3%) |
| Hypertension | 106 (30.6%) | 175 (36.5%) | 409 (23.8%) | 32 (33.3%) | 1,675 (28.0%) | 101 (33.7%) | 36 (20.2%) | 142 (33.9%) | 2,676 (27.4%) |
| Hyperlipidemia | 34 (9.8%) | 56 (11.7%) | 159 (9.3%) | 9 (9.4%) | 605 (10.1%) | 42 (14.0%) | 12 (6.7%) | 42 (10.0%) | 1,068 (10.9%) |
| Gerd | 20 (5.8%) | 21 (4.4%) | 57 (3.3%) | <5 | 238 (4.0%) | 12 (4.0%) | 11 (6.2%) | 14 (3.3%) | 379 (3.9%) |
| Cancer excl skin cancer | 56 (16.2%) | 40 (8.3%) | 211 (12.3%) | 14 (14.6%) | 854 (14.3%) | 39 (13.0%) | 9 (5.1%) | 47 (11.2%) | 1,070 (11.0%) |
| Obesity | 45 (13.0%) | 64 (13.3%) | 246 (14.3%) | 10 (10.4%) | 637 (10.7%) | 45 (15.0%) | 17 (9.6%) | 41 (9.8%) | 1,072 (11.0%) |
| Asthma | 51 (14.7%) | 62 (12.9%) | 188 (10.9%) | 12 (12.5%) | 636 (10.6%) | 38 (12.7%) | 20 (11.2%) | 56 (13.4%) | 1,116 (11.4%) |
| T 2 d | 39 (11.3%) | 73 (15.2%) | 173 (10.1%) | 11 (11.5%) | 652 (10.9%) | 50 (16.7%) | 13 (7.3%) | 39 (9.3%) | 1,070 (11.0%) |
| Anemia | 36 (10.4%) | 49 (10.2%) | 135 (7.9%) | 7 (7.3%) | 507 (8.5%) | 23 (7.7%) | 18 (10.1%) | 33 (7.9%) | 792 (8.1%) |
| Copd | 43 (12.4%) | 42 (8.8%) | 100 (5.8%) | 9 (9.4%) | 465 (7.8%) | 24 (8.0%) | <5 | 33 (7.9%) | 691 (7.1%) |
| Chronic kidney disease | 44 (12.7%) | 101 (21.0%) | 190 (11.1%) | 14 (14.6%) | 794 (13.3%) | 70 (23.3%) | 9 (5.1%) | 75 (17.9%) | 1,225 (12.5%) |
| Depressive disorder | 60 (17.3%) | 91 (19.0%) | 296 (17.2%) | 18 (18.8%) | 1,024 (17.1%) | 44 (14.7%) | 43 (24.2%) | 82 (19.6%) | 1,779 (18.2%) |
| Ischemic heart disease | 36 (10.4%) | 53 (11.0%) | 118 (6.9%) | 14 (14.6%) | 626 (10.5%) | 48 (16.0%) | 5 (2.8%) | 62 (14.8%) | 1,004 (10.3%) |
| Peripheral vascular disease | <5 | 7 (1.5%) | 26 (1.5%) | <5 | 121 (2.0%) | 9 (3.0%) | 0 (0.0%) | 9 (2.2%) | 191 (2.0%) |
| **Medications 365 days prior to index date (N%)** |  |  |  |  |  |  |  |  |  |
| Antiemetics | 6 (1.7%) | 5 (1.0%) | 17 (1.0%) | <5 | 29 (0.5%) | 0 (0.0%) | <5 | <5 | 57 (0.6%) |
| Systemic Antibacterials | 192 (55.5%) | 273 (56.9%) | 852 (49.6%) | 52 (54.2%) | 3,053 (51.1%) | 152 (50.7%) | 104 (58.4%) | 235 (56.1%) | 4,502 (46.1%) |
| Drugs for acid related disorders | 243 (70.2%) | 300 (62.5%) | 767 (44.6%) | 58 (60.4%) | 2,663 (44.5%) | 151 (50.3%) | 71 (39.9%) | 263 (62.8%) | 4,004 (41.0%) |
| Anti-inflammatories and antirheumatics | 209 (60.4%) | 277 (57.7%) | 814 (47.4%) | 64 (66.7%) | 1,970 (32.9%) | 142 (47.3%) | 64 (36.0%) | 247 (59.0%) | 2,943 (30.1%) |
| Antianaemic preparations | 101 (29.2%) | 103 (21.5%) | 304 (17.7%) | 35 (36.5%) | 1,211 (20.3%) | 44 (14.7%) | 39 (21.9%) | 77 (18.4%) | 1,594 (16.3%) |
| Diuretics | 96 (27.8%) | 157 (32.7%) | 337 (19.6%) | 24 (25.0%) | 1,495 (25.0%) | 119 (39.7%) | 20 (11.2%) | 132 (31.5%) | 2,217 (22.7%) |
| Psycholeptics | 106 (30.6%) | 121 (25.2%) | 372 (21.7%) | 31 (32.3%) | 1,264 (21.1%) | 55 (18.3%) | 43 (24.2%) | 99 (23.6%) | 1,772 (18.1%) |
| Antibiotics | 30 (8.7%) | 43 (9.0%) | 130 (7.6%) | 11 (11.5%) | 514 (8.6%) | 24 (8.0%) | 12 (6.7%) | 32 (7.6%) | 641 (6.6%) |
| Immunosupressants | 25 (7.2%) | 16 (3.3%) | 33 (1.9%) | 14 (14.6%) | 87 (1.5%) | <5 | <5 | 17 (4.1%) | 153 (1.6%) |
| Opioids | 224 (64.7%) | 242 (50.4%) | 740 (43.1%) | 56 (58.3%) | 2,380 (39.8%) | 119 (39.7%) | 58 (32.6%) | 212 (50.6%) | 3,667 (37.5%) |
| Calcium channel blockers | 70 (20.2%) | 141 (29.4%) | 313 (18.2%) | 22 (22.9%) | 1,368 (22.9%) | 87 (29.0%) | 19 (10.7%) | 109 (26.0%) | 2,120 (21.7%) |
| Beta blocking agents | 86 (24.9%) | 163 (34.0%) | 358 (20.8%) | 35 (36.5%) | 1,396 (23.3%) | 107 (35.7%) | 32 (18.0%) | 117 (27.9%) | 2,279 (23.3%) |
| Lipid modifying agents | 135 (39.0%) | 209 (43.5%) | 526 (30.6%) | 35 (36.5%) | 2,236 (37.4%) | 148 (49.3%) | 29 (16.3%) | 185 (44.2%) | 3,510 (35.9%) |
| Agents for renin-angiotensin-system | 121 (35.0%) | 218 (45.4%) | 471 (27.4%) | 37 (38.5%) | 2,009 (33.6%) | 152 (50.7%) | 31 (17.4%) | 160 (38.2%) | 3,340 (34.2%) |
| Antidepressants | 140 (40.5%) | 159 (33.1%) | 522 (30.4%) | 36 (37.5%) | 1,654 (27.7%) | 69 (23.0%) | 59 (33.2%) | 149 (35.6%) | 2,667 (27.3%) |
| Drugs used in diabetes | 44 (12.7%) | 62 (12.9%) | 185 (10.8%) | 11 (11.5%) | 704 (11.8%) | 42 (14.0%) | 12 (6.7%) | 50 (11.9%) | 1,109 (11.4%) |
| Drugs for addictive disorders | 12 (3.5%) | 11 (2.3%) | 58 (3.4%) | <5 | 182 (3.0%) | <5 | 6 (3.3%) | 9 (2.2%) | 282 (2.9%) |
| Hormonal contraceptives | 7 (2.0%) | 8 (1.7%) | 62 (3.6%) | <5 | 149 (2.5%) | <5 | 25 (14.0%) | 9 (2.2%) | 249 (2.6%) |
| Antithrombotics | 54 (15.6%) | 82 (17.1%) | 194 (11.3%) | 18 (18.8%) | 782 (13.1%) | 50 (16.7%) | 14 (7.9%) | 63 (15.0%) | 1,217 (12.5%) |
| Drugs for obstructive airway disorders | 122 (35.3%) | 167 (34.8%) | 469 (27.3%) | 33 (34.4%) | 1,614 (27.0%) | 90 (30.0%) | 44 (24.7%) | 128 (30.6%) | 2,604 (26.6%) |
| Antiepileptics | 77 (22.3%) | 70 (14.6%) | 204 (11.9%) | 14 (14.6%) | 629 (10.5%) | 34 (11.3%) | 20 (11.2%) | 58 (13.8%) | 945 (9.7%) |

### Table S4: Adjusted sequence ratios (ASRs) with 95% confidence intervals for cardiovascular events for negative control acetaminophen.

| **Index cohort name** | **Marker cohort name** | **Index N (%)** | **Marker N (%)** | **CSR (95% CI)** | **ASR (95% CI)** |
| --- | --- | --- | --- | --- | --- |
| Acetaminophen | Myocardial Infarction | 1,811 (30.0%) | 4,229 (70.0%) | 0.43 [0.41 - 0.45] | 0.33 [0.32 - 0.35] |
|  | Arrythmia | 6,457 (43.1%) | 8,533 (56.9%) | 0.76 [0.73 - 0.78] | 0.59 [0.57 - 0.61] |
|  | DVT | 2,211 (48.9%) | 2,308 (51.1%) | 0.96 [0.90 - 1.02] | 0.75 [0.71 - 0.80] |
|  | GI hemorrhage | 4,563 (49.9%) | 4,582 (50.1%) | 1.00 [0.96 - 1.04] | 0.79 [0.76 - 0.83] |
|  | Heart failure | 3,791 (48.0%) | 4,101 (52.0%) | 0.92 [0.88 - 0.97] | 0.73 [0.70 - 0.77] |
|  | Hemorrhagic stroke | 403 (23.1%) | 1,342 (76.9%) | 0.30 [0.27 - 0.34] | 0.23 [0.20 - 0.26] |
|  | Ischemic stroke | 2,225 (32.0%) | 4,739 (68.0%) | 0.47 [0.45 - 0.49] | 0.37 [0.35 - 0.38] |
|  | PE | 1,748 (41.7%) | 2,447 (58.3%) | 0.71 [0.67 - 0.76] | 0.55 [0.52 - 0.59] |
|  | Any stroke | 2,557 (30.2%) | 5,915 (69.8%) | 0.43 [0.41 - 0.45] | 0.34 [0.32 - 0.35] |

DVT; Deep vein thrombosis, GI; gastrointestinal, PE; pulmonary embolism, MI; Myocardial Infarction

### Table S5: Adjusted sequence ratios (ASRs), crude sequence ratios (CSR) with 95% confidence intervals for cardiovascular events and percentages of index and markers.

| Index cohort name | Marker cohort name | Index N (%) | Marker N (%) | CSR (95% CI) | ASR (95% CI) |
| --- | --- | --- | --- | --- | --- |
| Any NSAID | MI | 1,405 (73.8%) | 499 (26.2%) | 2.82 [2.54 - 3.12] | 2.32 [2.10 - 2.57] |
|  | Arrythmia | 3,948 (66.0%) | 2,036 (34.0%) | 1.94 [1.84 - 2.05] | 1.60 [1.52 - 1.69] |
|  | DVT | 1,825 (70.3%) | 770 (29.7%) | 2.37 [2.18 - 2.58] | 1.98 [1.82 - 2.15] |
|  | Heart failure | 1,229 (65.4%) | 649 (34.6%) | 1.89 [1.72 - 2.08] | 1.60 [1.46 - 1.76] |
|  | PE | 1,231 (75.0%) | 411 (25.0%) | 3.00 [2.68 - 3.35] | 2.44 [2.19 - 2.73] |
|  | Stroke | 1,537 (63.5%) | 884 (36.5%) | 1.74 [1.60 - 1.89] | 1.43 [1.32 - 1.56] |
| COX-2 inhibitor | MI | 83 (69.7%) | 36 (30.3%) | 2.31 [1.57 - 3.44] | 2.11 [1.44 - 3.15] |
|  | Arrythmia | 234 (61.1%) | 149 (38.9%) | 1.57 [1.28 - 1.93] | 1.43 [1.17 - 1.76] |
|  | DVT | 84 (64.6%) | 46 (35.4%) | 1.83 [1.28 - 2.63] | 1.68 [1.18 - 2.41] |
|  | Heart failure | 85 (70.2%) | 36 (29.8%) | 2.36 [1.61 - 3.51] | 2.18 [1.48 - 3.24] |
|  | PE | 71 (67.0%) | 35 (33.0%) | 2.03 [1.36 - 3.06] | 1.83 [1.23 - 2.77] |
|  | Stroke | 87 (73.7%) | 31 (26.3%) | 2.81 [1.88 - 4.27] | 2.56 [1.72 - 3.90] |
| Non selective | MI | 1,377 (73.4%) | 499 (26.6%) | 2.76 [2.49 - 3.06] | 2.28 [2.06 - 2.53] |
|  | Arrythmia | 3,853 (65.8%) | 2,001 (34.2%) | 1.93 [1.82 - 2.03] | 1.60 [1.51 - 1.69] |
|  | DVT | 1,797 (70.3%) | 759 (29.7%) | 2.37 [2.18 - 2.58] | 1.98 [1.82 - 2.16] |
|  | Heart failure | 1,187 (65.3%) | 632 (34.7%) | 1.88 [1.71 - 2.07] | 1.59 [1.45 - 1.75] |
|  | PE | 1,220 (75.4%) | 398 (24.6%) | 3.07 [2.74 - 3.44] | 2.51 [2.24 - 2.81] |
|  | Stroke | 1,510 (63.3%) | 877 (36.7%) | 1.72 [1.58 - 1.87] | 1.42 [1.31 - 1.55] |
| Aspirin | MI | 2,595 (8.1%) | 29,533 (91.9%) | 0.09 [0.08 - 0.09] | 0.07 [0.07 - 0.07] |
|  | Arrythmia | 6,429 (43.8%) | 8,241 (56.2%) | 0.78 [0.76 - 0.81] | 0.63 [0.61 - 0.65] |
|  | DVT | 647 (48.4%) | 691 (51.6%) | 0.94 [0.84 - 1.04] | 0.76 [0.69 - 0.85] |
|  | Heart failure | 3,735 (40.5%) | 5,482 (59.5%) | 0.68 [0.65 - 0.71] | 0.56 [0.54 - 0.59] |
|  | PE | 608 (57.4%) | 452 (42.6%) | 1.35 [1.19 - 1.52] | 1.08 [0.96 - 1.22] |
|  | Stroke | 2,848 (37.6%) | 4,727 (62.4%) | 0.60 [0.58 - 0.63] | 0.49 [0.47 - 0.51] |
| Celecoxib | MI | 30 (66.7%) | 15 (33.3%) | 2.00 [1.09 - 3.78] | 1.88 [1.02 - 3.55] |
|  | Arrythmia | 100 (62.9%) | 59 (37.1%) | 1.69 [1.23 - 2.35] | 1.59 [1.15 - 2.19] |
|  | DVT | 41 (67.2%) | 20 (32.8%) | 2.05 [1.21 - 3.54] | 1.93 [1.14 - 3.34] |
|  | Heart failure | 32 (69.6%) | 14 (30.4%) | 2.29 [1.24 - 4.37] | 2.15 [1.17 - 4.11] |
|  | PE | 36 (62.1%) | 22 (37.9%) | 1.64 [0.97 - 2.81] | 1.52 [0.90 - 2.60] |
|  | Stroke | 33 (67.3%) | 16 (32.7%) | 2.06 [1.15 - 3.81] | 1.93 [1.08 - 3.56] |
| Diclofenac | MI | 210 (82.0%) | 46 (18.0%) | 4.57 [3.34 - 6.33] | 3.30 [2.42 - 4.57] |
|  | Arrythmia | 473 (66.7%) | 236 (33.3%) | 2.00 [1.72 - 2.35] | 1.44 [1.23 - 1.68] |
|  | DVT | 255 (65.9%) | 132 (34.1%) | 1.93 [1.57 - 2.39] | 1.40 [1.14 - 1.73] |
|  | Heart failure | 107 (65.6%) | 56 (34.4%) | 1.91 [1.39 - 2.65] | 1.40 [1.02 - 1.94] |
|  | PE | 204 (73.4%) | 74 (26.6%) | 2.76 [2.12 - 3.61] | 1.99 [1.53 - 2.60] |
|  | Stroke | 198 (69.5%) | 87 (30.5%) | 2.28 [1.77 - 2.94] | 1.64 [1.28 - 2.12] |
| Etodolac | Arrythmia | 39 (67.2%) | 19 (32.8%) | 2.05 [1.20 - 3.60] | 1.74 [1.02 - 3.05] |
|  | DVT | 7 (50.0%) | 7 (50.0%) | 1.00 [0.35 - 2.86] | 0.86 [0.30 - 2.46] |
|  | Heart failure | 8 (53.3%) | 7 (46.7%) | 1.14 [0.42 - 3.19] | 0.99 [0.36 - 2.77] |
|  | Stroke | 16 (76.2%) | 5 (23.8%) | 3.20 [1.24 - 9.29] | 2.72 [1.06 - 7.89] |
| Etoricoxib | MI | 55 (71.4%) | 22 (28.6%) | 2.50 [1.54 - 4.15] | 2.27 [1.40 - 3.76] |
|  | Arrythmia | 142 (59.7%) | 96 (40.3%) | 1.48 [1.14 - 1.92] | 1.34 [1.04 - 1.74] |
|  | DVT | 45 (63.4%) | 26 (36.6%) | 1.73 [1.08 - 2.83] | 1.58 [0.98 - 2.58] |
|  | Heart failure | 55 (70.5%) | 23 (29.5%) | 2.39 [1.49 - 3.94] | 2.19 [1.36 - 3.61] |
|  | PE | 39 (75.0%) | 13 (25.0%) | 3.00 [1.64 - 5.76] | 2.69 [1.47 - 5.15] |
|  | Stroke | 61 (80.3%) | 15 (19.7%) | 4.07 [2.37 - 7.33] | 3.68 [2.14 - 6.62] |
| Ibuprofen | MI | 514 (69.5%) | 226 (30.5%) | 2.27 [1.95 - 2.66] | 1.92 [1.65 - 2.25] |
|  | Arrythmia | 1,501 (64.0%) | 846 (36.0%) | 1.77 [1.63 - 1.93] | 1.51 [1.38 - 1.64] |
|  | DVT | 776 (69.0%) | 348 (31.0%) | 2.23 [1.97 - 2.53] | 1.91 [1.68 - 2.17] |
|  | Heart failure | 540 (68.0%) | 254 (32.0%) | 2.13 [1.83 - 2.47] | 1.84 [1.59 - 2.14] |
|  | PE | 553 (72.6%) | 209 (27.4%) | 2.65 [2.26 - 3.11] | 2.20 [1.88 - 2.59] |
|  | Stroke | 625 (58.9%) | 436 (41.1%) | 1.43 [1.27 - 1.62] | 1.21 [1.07 - 1.37] |
| Indomethacin | MI | 28 (65.1%) | 15 (34.9%) | 1.87 [1.01 - 3.55] | 1.46 [0.79 - 2.77] |
|  | Arrythmia | 82 (52.6%) | 74 (47.4%) | 1.11 [0.81 - 1.52] | 0.87 [0.63 - 1.19] |
|  | DVT | 25 (62.5%) | 15 (37.5%) | 1.67 [0.89 - 3.20] | 1.31 [0.70 - 2.53] |
|  | Heart failure | 38 (58.5%) | 27 (41.5%) | 1.41 [0.86 - 2.32] | 1.13 [0.69 - 1.85] |
|  | PE | 14 (63.6%) | 8 (36.4%) | 1.75 [0.75 - 4.28] | 1.36 [0.58 - 3.33] |
|  | Stroke | 34 (65.4%) | 18 (34.6%) | 1.89 [1.08 - 3.39] | 1.47 [0.84 - 2.64] |
| Mefenamate | Arrythmia | 33 (50.0%) | 33 (50.0%) | 1.00 [0.62 - 1.62] | 0.88 [0.54 - 1.43] |
|  | DVT | 29 (54.7%) | 24 (45.3%) | 1.21 [0.71 - 2.08] | 1.07 [0.63 - 1.85] |
|  | Heart failure | 8 (61.5%) | 5 (38.5%) | 1.60 [0.54 - 5.07] | 1.44 [0.48 - 4.55] |
|  | PE | 20 (54.1%) | 17 (45.9%) | 1.18 [0.62 - 2.26] | 1.02 [0.54 - 1.97] |
|  | Stroke | 16 (64.0%) | 9 (36.0%) | 1.78 [0.80 - 4.12] | 1.56 [0.71 - 3.62] |
| Meloxicam | MI | 43 (68.3%) | 20 (31.7%) | 2.15 [1.28 - 3.70] | 1.80 [1.07 - 3.09] |
|  | Arrythmia | 139 (69.5%) | 61 (30.5%) | 2.28 [1.69 - 3.09] | 1.90 [1.41 - 2.58] |
|  | DVT | 36 (58.1%) | 26 (41.9%) | 1.38 [0.84 - 2.31] | 1.16 [0.71 - 1.94] |
|  | Heart failure | 38 (62.3%) | 23 (37.7%) | 1.65 [0.99 - 2.80] | 1.40 [0.84 - 2.37] |
|  | PE | 29 (76.3%) | 9 (23.7%) | 3.22 [1.58 - 7.05] | 2.66 [1.30 - 5.82] |
|  | Stroke | 56 (68.3%) | 26 (31.7%) | 2.15 [1.37 - 3.46] | 1.79 [1.14 - 2.88] |
| Naproxen | MI | 964 (71.8%) | 379 (28.2%) | 2.54 [2.26 - 2.87] | 2.34 [2.08 - 2.64] |
|  | Arrythmia | 2,677 (65.4%) | 1,415 (34.6%) | 1.89 [1.77 - 2.02] | 1.75 [1.64 - 1.87] |
|  | DVT | 1,251 (70.6%) | 521 (29.4%) | 2.40 [2.17 - 2.66] | 2.24 [2.02 - 2.48] |
|  | Heart failure | 779 (64.8%) | 423 (35.2%) | 1.84 [1.64 - 2.07] | 1.74 [1.54 - 1.96] |
|  | PE | 795 (77.0%) | 237 (23.0%) | 3.35 [2.91 - 3.88] | 3.03 [2.63 - 3.51] |
|  | Stroke | 1,057 (64.9%) | 572 (35.1%) | 1.85 [1.67 - 2.05] | 1.70 [1.54 - 1.88] |

DVT; Deep vein thrombosis, GI; gastrointestinal, PE; pulmonary embolism, MI; Myocardial Infarction

### Figure S1: Adjusted sequence ratios (ASRs) with 95% confidence intervals for hemorrhage stroke and ischemic stroke.

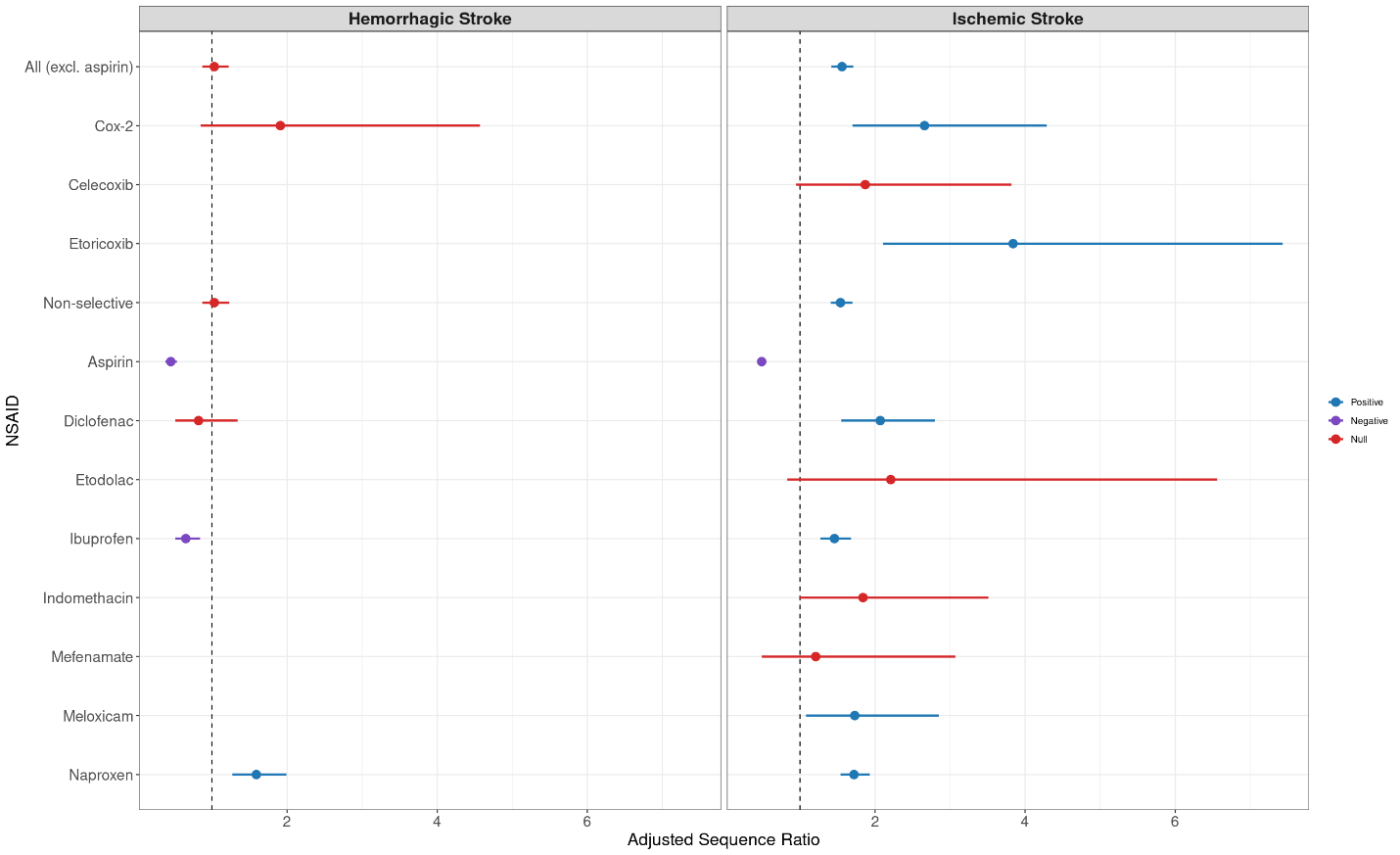

### Figure S2: Sex stratified adjusted sequence ratios (ASRs) with 95% confidence intervals for cardiovascular events.

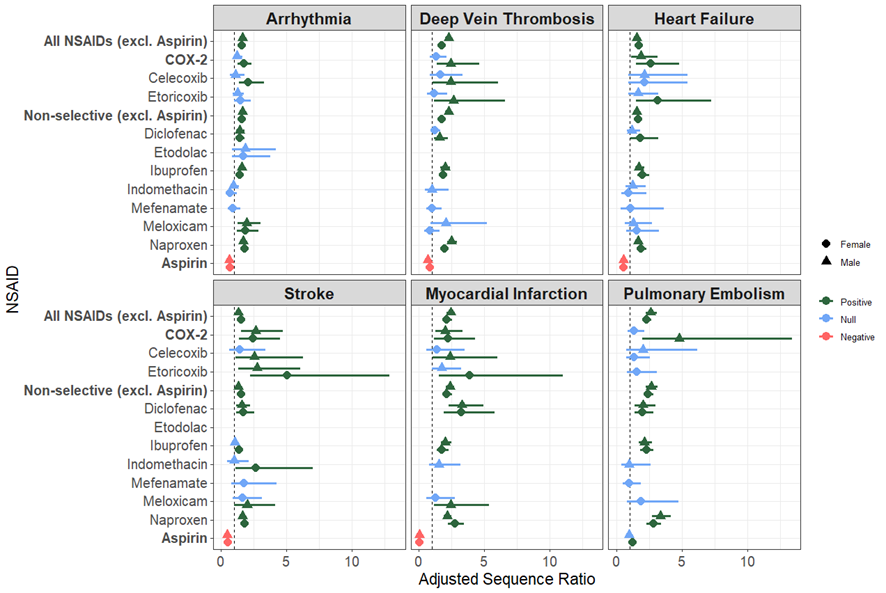

### Figure S3: Age stratified adjusted sequence ratios (ASRs) with 95% confidence intervals for cardiovascular events.

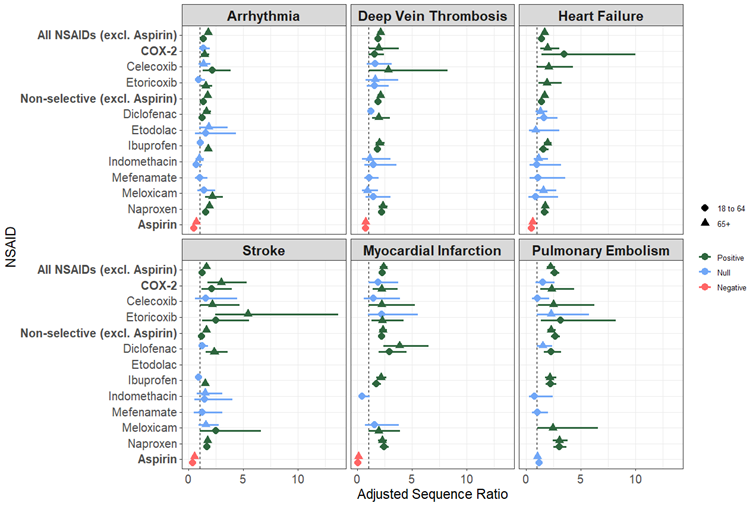

### Figure S4: Adjusted sequence ratios (ASRs) with 95% confidence intervals for cardiovascular events within a 90-day initiation window.

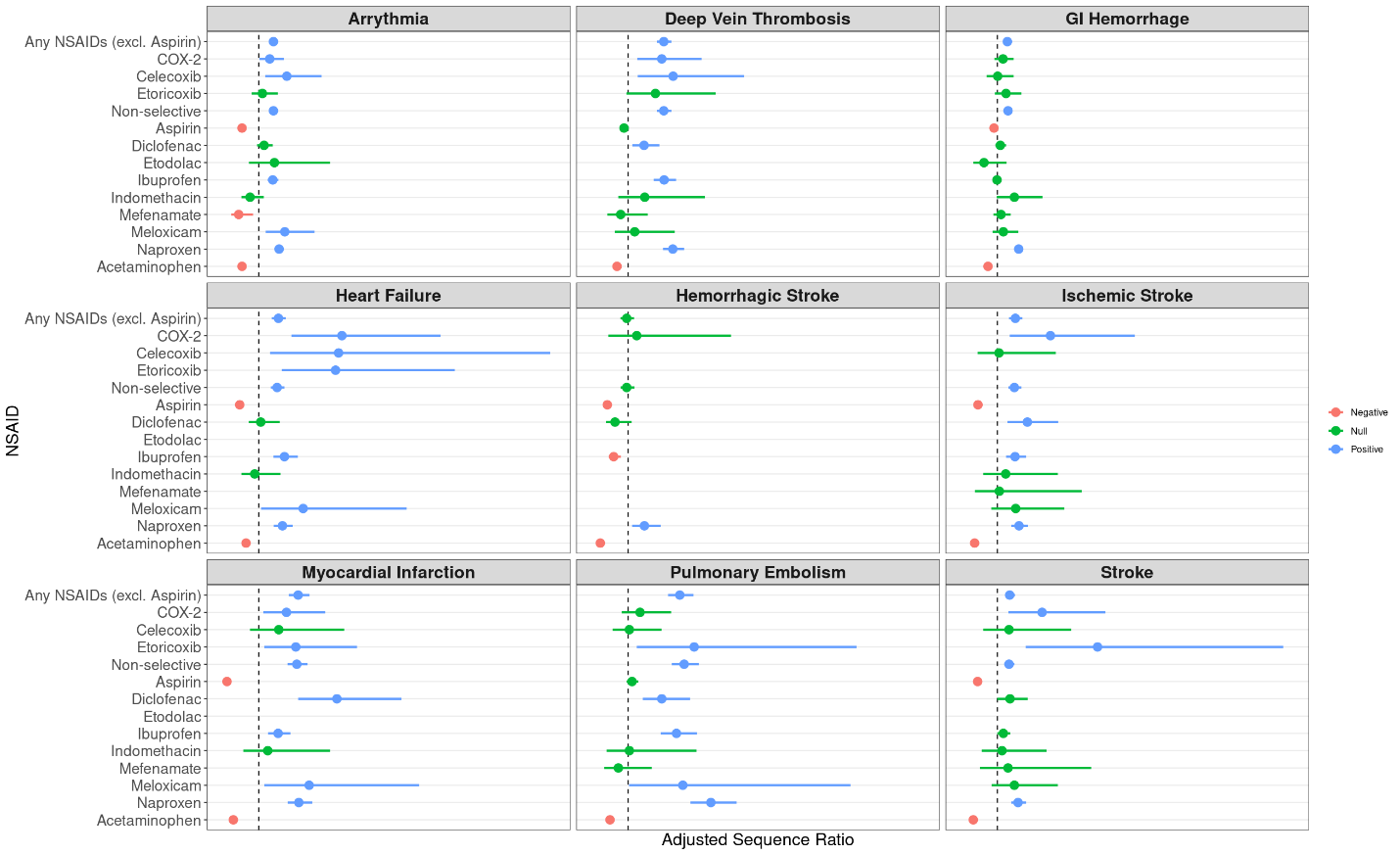

### Figure S5: Adjusted sequence ratios (ASRs) with 95% confidence intervals for cardiovascular events within a 365-day initiation window.

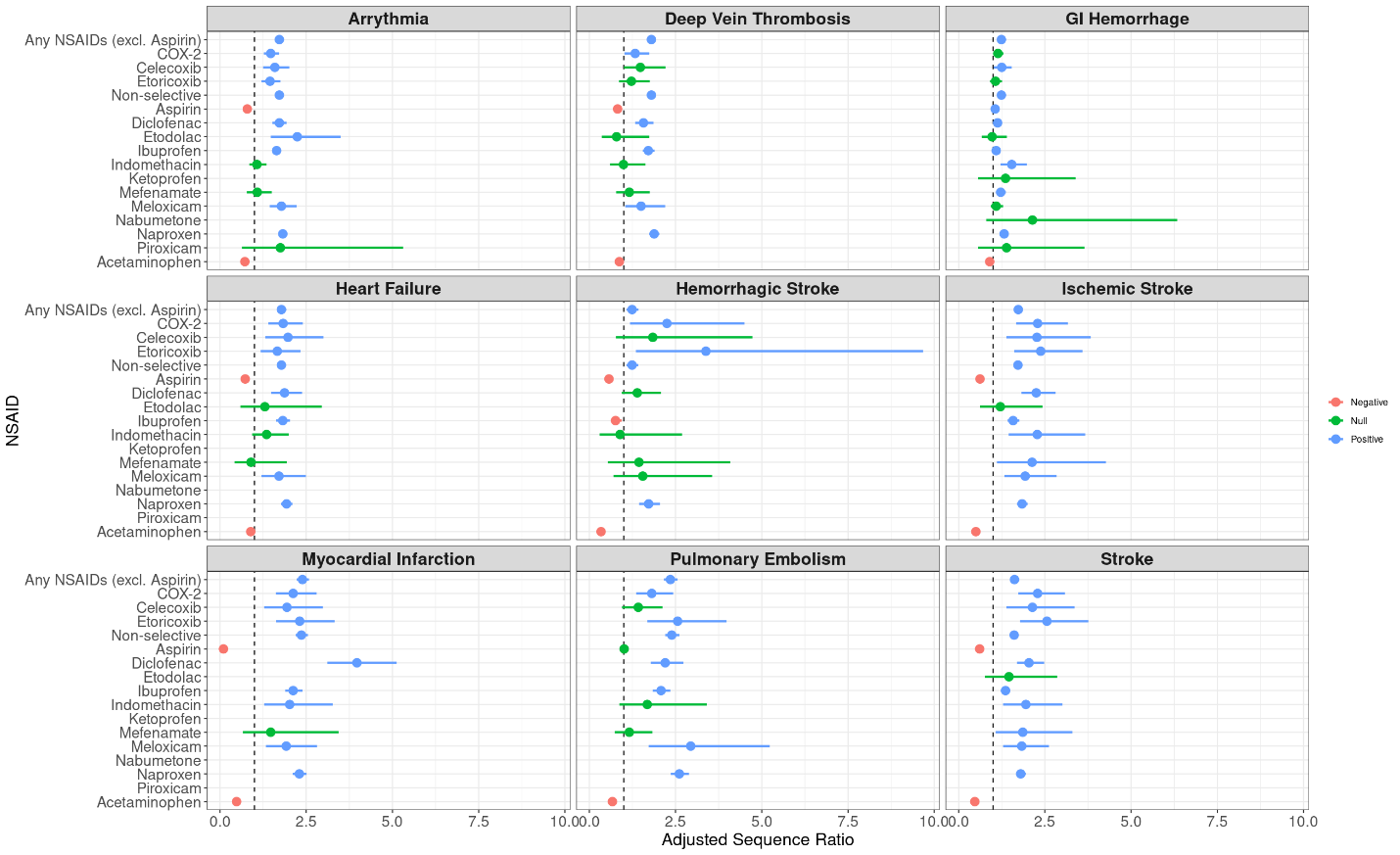

### Figure S6: Adjusted sequence ratios (ASRs) with 95% confidence intervals for cardiovascular events stratified by prior PPI use for any NSAIDs.

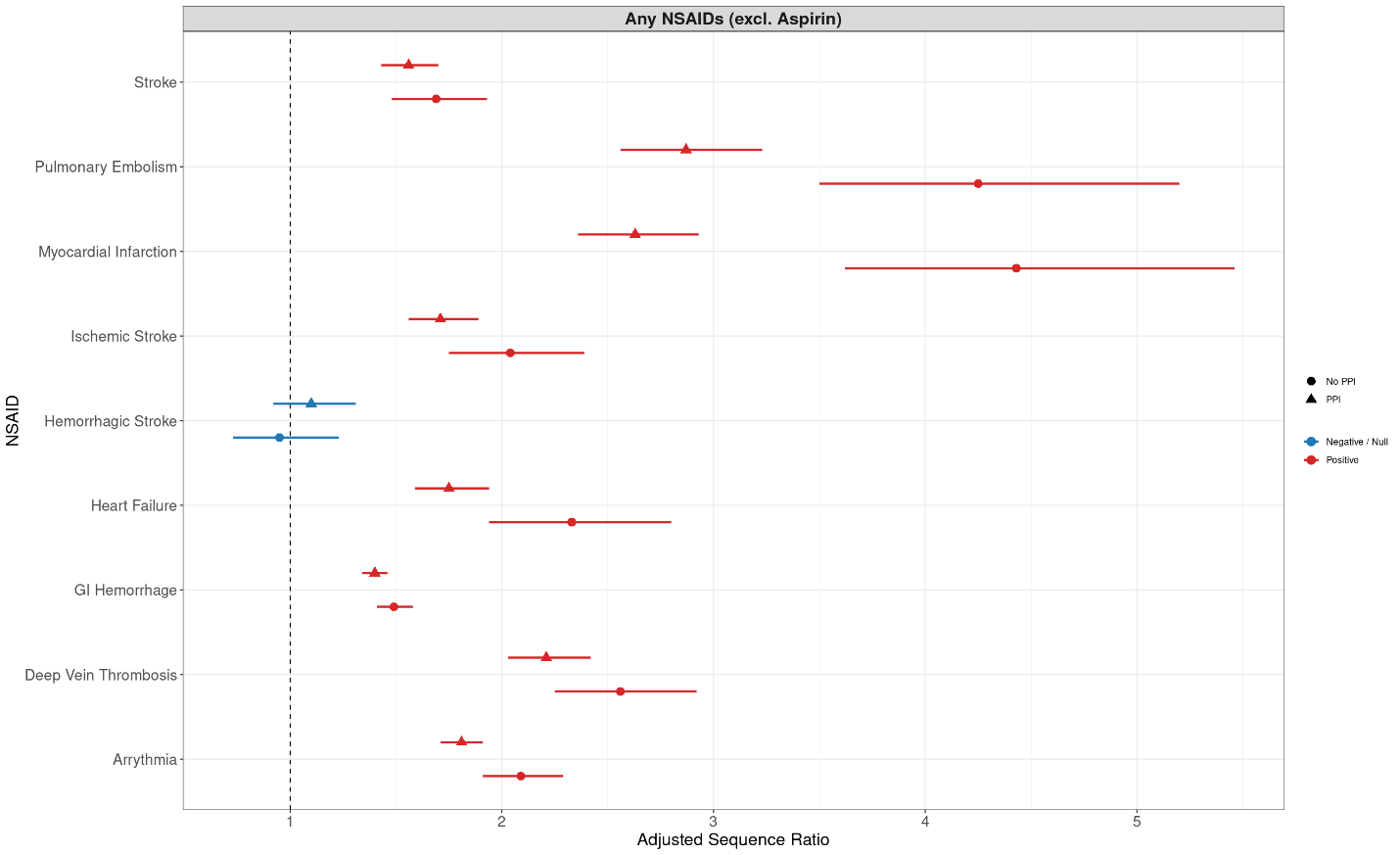
